# Location patterns and longitudinal progression of white matter hyperintensities

**DOI:** 10.64898/2026.02.20.26346709

**Authors:** Xin Zhao, Ian B. Malone, Thomas M. Brown, Andrew Wong, David M. Cash, Nish Chaturvedi, Alun D. Hughes, Jonathan M. Schott, Frederik Barkhof, Josephine Barnes, Carole H. Sudre, Alzheimer’s Disease Neuroimaging Initiative

## Abstract

**Background and Objectives:** White matter hyperintensities (WMH) of presumed vascular origin are a neuroimaging hallmark of cerebral small vessel disease (CSVD). Their spatial heterogeneity may reflect different clinical phenotypes. Most prior studies relied on principal component analysis to characterise such heterogeneity, which has limited ability to stratify individuals into discrete and interpretable WMH subtypes. We therefore propose a data-driven framework to identify WMH spatial subtypes, characterise their demographic and clinical profiles, and investigate their predictive value for future WMH progression.

**Methods:** We analysed MRI scans from 63,338 individuals across 4 major cohorts (internal data): ADNI3, Insight46, SABRE and UK Biobank (UKB), and validated our findings in the OASIS-3 dataset (n=844). WMH were automatically segmented and regionally quantified using a 36-region bullseye framework. Clustering was applied to the relative regional distributions of WMH. A stability-based approach was used to identify robust WMH subtypes. Their associations with 19 risk factors of interest were analysed using multivariable regression. In a subset with follow-up MRI scans (internal: n=5,274, OASIS-3: n=182), we evaluated the predictive value of these subtypes combined with other volumetric or spatial WMH variables for WMH progression.

**Results:** Five WMH location patterns with different lesion burden and spatial distribution were identified (stability score 0.946) and reproduced in OASIS-3. These patterns showed distinct associations with demographic, vascular, metabolic, inflammatory and genetic risk factors. Higher-burden patterns were independently associated with older age, higher blood pressure, diabetes and smoking, indicating a gradient of vascular risk across spatial subtypes. WMH location patterns were largely preserved over 18-30 months, with most individuals remaining within the same pattern (71.5%). While global baseline WMH volume remained a strong predictor of future WMH progression (balanced accuracy 0.693, 95% CI: 0.664-0.723), models including baseline regional WMH volumes consistently outperformed other candidates (best balanced accuracy 0.737, 95% CI: 0.706-0.764).

**Discussion:** We presented a robust and scalable framework for spatial WMH phenotyping. We discussed clinical and prognostic implications of the spatial subtypes beyond total lesion burden. Our findings supported the value of WMH spatial characterisation in stratifying risk that may help guide personalised approaches to managing CSVD.

## 1 Introduction

White matter hyperintensities (WMH) of presumed vascular origin are a common neuroimaging manifestation of cerebral small vessel disease (CSVD) associated with ageing, dementia, stroke, and cognitive decline [1], [2], with emerging evidence suggesting both vascular and neurodegenerative contributions [3]. These hyperintense regions on magnetic resonance imaging (MRI), visible on T2-weighted images such as fluid-attenuated inversion recovery (T2-FLAIR), reflect increased water content and structural damage within white matter [4] with demyelination occurring in more severe cases. WMH prevalence increases with age, observed in 20-50% of middle-aged populations and over 90% of the elderly population [2]. In addition to age, WMH are also influenced by cardiovascular risk factors and genetic predispositions, contributing to heterogeneity across populations [2].

WMH are not randomly distributed across the brain, and their spatial heterogeneity correlates with clinical characteristics [5], [6], [7]. WMH burden in strategic white matter tracts or regions is associated with processing speed and executive function [5], and dementia risk [8] than global WMH burden alone. Regional WMH patterns [8], [9], [10] also show stronger associations with vascular risk factors and pathology. Additionally, the rate and pattern of WMH progression vary regionally, with periventricular and frontal lesions progressing more rapidly over time [6], [11].

Recent studies have shown increasing interest in WMH localisation beyond traditional regional divisions (*i.e.* periventricular vs deep WMH, or lobar WMH). A bullseye method [12] was proposed to address limitations of coarse regional characterisation. This framework offers a standardised, detailed and patient-specific regional representation of WMH burden across cortical and subcortical areas, represented by a bullseye. Existing work has applied principal component analysis (PCA) to regional or lobar WMH measures [10], [13], [14], [15]. PCA was used to capture shared covariance between regions and highlight spatial configurations linked to cognitive outcomes. However, it did not capture distinct WMH presentations and stratify individuals into discrete subtypes.

Both manual and data-driven methods have been proposed to capture individual spatial variation in WMH. A visual classification scheme grouped WMH into four spatial patterns – multi-spots, peri-basal ganglia, anterior subcortical patches and posterior subcortical patches based on expert rating in an ageing cohort [7]. Such visual methods require expert input and do not scale. More recently, unsupervised data-driven approaches have emerged. For instance, voxel-level spectral clustering identified five spatial signatures (juxtacortical, deep frontal, PV, parietal, and posterior WMH) associated with distinct CSVD aetiologies in the Alzheimer’s Disease Neuroimaging Initiative (ADNI) [16]. A multi-centre study adopted an unsupervised anomaly detection approach and provided a voxel-wise WMH distribution map across 11 memory-clinic cohorts (including ADNI) [17].

Despite this progress, discrete classification of spatial WMH subtypes remains limited. We therefore proposed and evaluated a robust data-driven framework to identify spatial WMH subtypes and investigated their relationship with risk factors and disease evolution. Clustering with stability selection was used to capture distinct and reproducible spatial patterns. To examine their clinical relevance, we examined cross-sectional associations with demographic, clinical, and genetic factors. To assess longitudinal relevance, we evaluated the prognostic value of baseline spatial WMH features for predicting WMH progression. Overall, our study aimed to provide a comprehensive characterisation of WMH location patterns and support patient stratification.

## 2 Methods

### 2.1 Data and processing

#### 2.1.1 Multi-cohort dataset

We used data from four studies for internal training and validation: the Alzheimer’s Disease Neuroimaging Initiative 3 (ADNI3), Insight46, the Southall and Brent Revisited (SABRE) cohort study, and the UK Biobank (UKB). We used the Open Access Series of Imaging Studies 3 (OASIS-3) as the validation dataset. Access and permission to use these data were granted prior to this study. All withdrawn participants were excluded. We included participants only if they had usable T1-weighted and FLAIR scans at one or more time points. Dataset descriptions are provided in Section e1.1, MRI protocols are summarised in eTable 1.

**ADNI3**^1^ is a multi-centre, prospective study in North America [18], [19], [20]. Participants were aged 55-90 at baseline, with no significant neurological disease other than Alzheimer’s disease.

**Insight46** Insight46 is the prospective neuroimaging substudy of the MRC National Survey for Health and Disease (NSHD) birth cohort [21], [22]. Participants were all born in the same week of March 1946. Participants were imaged in late midlife, with baseline imaging at ages 69-71 and follow-up data at ages 73-75.

**SABRE** The SABRE cohort is a longitudinal study based in West London [23], originally established to investigate cardiovascular disease and diabetes in people of White British, first-generation migrants of South Asian or African Caribbean heritage. Participants were aged 40-69 at baseline. Neuroimaging data were acquired in the third follow-up wave in mid-to late adulthood.

**UKB** UKB is a large population-based biomedical cohort in the United Kingdom (UK). UKB recruited participants aged 40-69 between 2006 and 2010 [24]. A subset of approximately 67,000 participants underwent brain MRI scans [25] with follow-ups available in some.

##### Validation dataset

**OASIS-3** OASIS-3 is a single-site longitudinal retrospective neuroimaging cohort from the United States (US) aged 42-95 [26]. It includes cognitively normal participants and those at various stages of cognitive impairment and dementia.

#### 2.1.2 Standard protocol approvals, registrations, and patient consents

All ADNI3 participants provided written informed consent under the institutional review boards of all participating sites. Insight46 was approved by the Health Research Authority Research Ethics Committee (HRA REC) London (reference 14/LO/1173) [22]. All participants provided their informed written consent at the beginning of each of the three phases, including permission for their data to be stored in accordance with the Data Protection Act (2018) and General Data Protection Regulations (GDPR). Ethical approval for SABRE investigations was obtained from the Fulham Research Ethics Committee (reference 14/LO/0108), and all participants gave written informed consent. UKB has received ethical approval from the National Health Service North West Centre for Research Ethics Committee (reference: 11/NW/0382). All participants provided consent during data collection to utilise their anonymous data. All OASIS-3 participants provided informed consent under procedures approved by the institution’s institutional review board.

#### 2.1.3 Volumetric quantification of WMH

WMH masks were segmented using the publicly available BaMoS [27] and binarised using a probability threshold of 0.5 (WMH vs non-WMH). WMH volumes were computed using true voxel dimensions to account for non-isotropic resolution, using NiftySeg^2^ software. Intracranial volume (ICV) was estimated from Geodesic Information Flows (GIF) [28] parcellation outputs. While WMH segmentations in Insight46 [21] and SABRE [29] underwent rigorous in-house quality control (QC), applying such procedures across the larger ADNI3, UKB and OASIS-3 datasets was not tractable due to scale. Therefore, we adopted a targeted QC approach for these cohorts (Section e1.2).

WMH masks were divided into 36 brain regions using cortical parcellation based on the Laplacian distance maps, and mapped using the bullseye representation [12]. The regions come from 9 lobes (*i.e.* left/right frontal, left/right parietal, left/right occipital, left/right temporal, and basal ganglia, thalamus and infratentorial^3^) and 4 equidistant layers.

#### 2.1.4 Normalisation of WMH volumes and preparation of distribution markers

To account for inter-individual variability relative to head size, total and regional WMH volumes were normalised by ICV (Equation 1). This approach is widely adopted and less affected by brain atrophy than normalisation using brain volumes [14], [30], [31], [32]. Unless otherwise stated, these ICV-normalised measures are referred to as WMH volumes below. For visualisation and reporting in volumetric units, however, they were additionally scaled to a common reference using the median baseline ICV from the internal dataset.

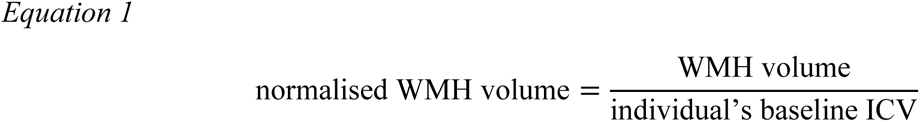

The relative distribution of WMH across regions was calculated by dividing regional volumes by total WMH volume, with 36 regional proportions summing to 1.

#### 2.1.5 Non-imaging variables

We analysed 19 non-imaging variables of interest based on existing findings on WMH. These included age [2], [30], [33], [34], sex [35] (male/female), ethnicity (white/black/Asian/mixed/other)^4^, systolic [30], [35] and diastolic blood pressures [35] (SBP and DBP), smoking status [30], [35] (never/ever/current), Type 2 diabetes diagnosis [30], [35] (no/yes), hypertension (no/yes) [2], use of lipid-lowering (MedChol), diabetic (MedDM) or anti-hypertensive medication (MedBP) [36] (no/yes). We also included a set of cardiometabolic and systemic blood markers: glucose [30], HbA1c [31], high-density lipoprotein cholesterol (HDL), triglycerides, low-density lipoprotein cholesterol (LDL) [2], white blood cell count (WBCC), and creatinine [37]. Lastly, APOE4 genotype [38] (0/1/2 alleles)^5^ was included. Availability of variables across cohorts are summarised in eTable 2.

### 2.2 Clustering and identification of location patterns

#### 2.2.1 Clustering methods

We compared clustering methods from three families: (1) k-means-based approaches, including standard k-means^6^ [39], subKmeans [40], which jointly learns an optimal clustering subspace and partition, and mini-batch k-means^7^; (2) a probabilistic Gaussian Mixture Model (GMM)^8^; (3) neural network-based methods, including Deep Embedded Clustering (DEC) [41], [42] and Deep k-Means (DKM) [43], which jointly learn latent representations and clustering.

#### 2.2.2 Stability evaluation using Jaccard index

Jaccard index-based stability [44] was used to quantify the consistency of clustering assignments across bootstrapped and complete-data iterations. Stability scores ranged between 0 (no overlap in cluster assignments) and 1 (identical cluster assignments).

#### 2.2.3 Stability selection for optimal *k*

Clustering was applied to relative regional distributions of WMH across 36 regions (summing to 1), following square-root-transformation and min-max scaling. A bootstrap-based stability estimation method [44] was adopted to identify the optimal *k* in the internal dataset. At each *k*, pairwise co-memberships across all bootstrapped and complete-data clusterings were evaluated using Jaccard index. The evaluation was carried out on the complete set of observations to allow for consistent comparison in stability across bootstrap iterations, without relying on direct centroid mapping. The latter could become unreliable when clusters split or merge across resamples. At each *k*, we evaluated the stability using a procedure analogous to 5-fold cross-validation (Section e1.7). From there, the most stable clustering was identified, and its average global Jaccard-based stability scores were used to construct a stability profile over a given range of *k*.

The optimal number of clusters *k* was selected as the maximum value between 2 and 7 for which the average Jaccard-based stability over 20 bootstrap iterations exceeded the recommended threshold of 0.90 for well-separated clusters [44]. Higher values of *k* were not considered due to reduced interpretability of clusters. Centroid estimation was obtained from all observations with stability greater than 0.90.

### 2.3 Statistical analyses

#### Cohort-wise participant characteristics

We compared participant characteristics between the internal and OASIS-3 datasets using Mann-Whitney U tests for continuous variables and Chi-squared tests for categorical variables.

#### Cross-sectional association of risk factors and WMH location patterns

We examined the cross-sectional associations of WMH location patterns (*i.e.* the clusters) with 19 non-imaging risk factors using multivariable linear regressions in the internal dataset. For each participant, Euclidean distances to cluster centroids were calculated in the clustering feature space, and proximity (inverse distance) was used as the outcome. Each model assessed the effects of one risk factor on the proximity to the centroid of one location pattern, adjusted for age, sex and baseline total WMH volume. All continuous variables were normalised to [0,1], and categorical variables were encoded as binary indicators. P-values were adjusted using Benjamini-Hochberg (BH) procedure [45]. The procedure was repeated in OASIS-3 for available variables. Missing variables were handled using complete-case analysis, assuming data were missing at random (also applies to analyses below).

#### Comparing risk factors and location patterns between WMH progression groups

Longitudinal analyses were conducted in participants with both baseline and follow-up MRI scans from ADNI3, Insight46 and UKB (follow-up interval of 1-7 years). We examined associations of 19 non-imaging risk factors, total baseline WMH volume, and distance variables with WMH progression in the internal dataset. We computed the annualised change in total WMH volume. A threshold equivalent to 250 *mm*^3^/year [36], [37], [46] (normalised WMH scale) was used to dichotomise participants into progressors and non-progressors. Between-group differences were assessed using Mann-Whitney U and Chi-squared tests, after removing the effect of age and sex. P-values were adjusted using BH procedure [44]. The above procedure was repeated in OASIS-3 in the eligible longitudinal subset.

### 2.4 Longitudinal prediction of WMH progression

#### Data and outcome variable

We used the longitudinal subset described in Section 2.3. The primary outcome was the binary threshold-based progression status.

#### Predictors

We included baseline predictors: risk factors (age, sex and systolic blood pressure), total and regional WMH volumes, Euclidean distances to cluster centroids, and principal component (PC) scores derived from regional WMH volumes. We fitted PCA on the training set and retained PCs explaining more than 80% variance. The test data was transformed using the training-derived parameters.

#### Model training

An XGBoost classifier [47] was trained using a random 80/20 train-test split. Hyperparameters were tuned (number of estimators: 50, 100, 200, maximum tree depth: 2, 3, 4 and 5) using 5-fold cross-validation on the training set, with inverse class weighting to address class imbalance. Models were selected and refit based on cross-validated balanced accuracy and evaluated on the internal test set and OASIS-3.

Performance was assessed using balanced accuracy^9^, which is more robust to class imbalance [48]. Training performance was summarised across cross-validation folds, test and OASIS-3 performance was estimated via bootstrap resampling (Section e1.8). We used McNemar’s test with BH-correction [45] for paired model comparison.

### 2.5 Sensitivity analyses

Sensitivity analyses assessed the robustness of the identified WMH location patterns. First, clustering was repeated after stratifying participants by baseline WMH volume (0-4mL and 4-8mL) and within a restricted age group (67-72 years). Second, longitudinal observations were conducted among participants with a follow-up interval of 18-30 months. Subtype transition frequencies and their stability at baseline were reviewed, alongside Euclidean distances to baseline-derived cluster centroids at baseline and follow-up.

## 3 Results

### 3.1 Participant characteristics

After applying exclusion criteria, our study included 1,007 participants from ADNI3, 462 participants from Insight46, 765 participants from SABRE, and 61,104 participants from UK Biobank. The validation set included 844 participants from OASIS-3. Participant flowcharts are provided in Section e2.1. The baseline characteristics of the internal dataset (n=63,338) and OASIS-3 (n=844) are presented in Table 1. Participants in OASIS-3 were approximately five years older (median: 71.3 [IQR: 65.9, 76.2]) than the internal dataset (66.2 [59.9, 71.5]), which aligned with their higher WMH burden (2.5 [1.3, 5.8]mL vs 1.8 [1.1, 3.9]mL).

**Table 1.** Baseline characteristics of internal dataset and OASIS-3.

|  | CATEGORY | UNIT | INTERNAL (N=63,338) | OASIS-3 (N=844) | P-VALUE |
| --- | --- | --- | --- | --- | --- |
| Age<sup>a</sup> |  | N<sup>b</sup> | 63,026 | 844 | < 0.001 |
|  |  | years | 66.2 [59.9, 71.5] | 71.3 [65.9, 76.2] |  |
| Sex<sup>c</sup> |  | N | 63,289 | 844 | < 0.001 |
|  | Female | n (%) | 33,401 (52.8%) | 488 (66.9%) |  |
|  | Male | n (%) | 29,888 (47.2%) | 241 (33.1%) |  |
| Ethnicity |  | N | 62,646 | 844 | n/a<sup>e</sup> |
|  | Asian | n (%) | 1,137 (1.8%) | < 10<sup>d</sup> |  |
|  | Black | n (%) | 698 (1.1%) | 106 (14.4%) |  |
|  | Mixed | n (%) | 342 (0.5%) | < 10<sup>d</sup> |  |
|  | Other | n (%) | 354 (0.6%) | – |  |
|  | White | n (%) | 60,115 (96.0%) | 622 (84.5%) |  |
| Total baseline WMH volume |  | N | 63,338 | 844 | < 0.001 |
|  |  | mL | 1.8 [1.1, 3.9] | 2.5 [1.3, 5.8] |  |
| APOE4 |  | N | 59,292 | 844 | < 0.001 |
|  | None | n (%) | 42,957 (72.4%) | 437 (59.7%) |  |
|  | 1 allele | n (%) | 15,023 (25.3%) | 254 (34.7%) |  |
|  | 2 alleles | n (%) | 1,312 (2.2%) | 41 (5.6%) |  |
| Smoking |  | N | 61,529 | 844 | < 0.001 |
|  | Current | n (%) | 2,061 (3.3%) | 36 (6.4%) |  |
|  | Ever | n (%) | 20,880 (33.9%) | 226 (39.9%) |  |
|  | Never | n (%) | 38,588 (62.7%) | 304 (53.7%) |  |
| Hypertension |  | N | 62,849 | 844 | < 0.001 |
|  | No | n (%) | 43,749 (69.6%) | 239 (53.0%) |  |
|  | Yes | n (%) | 19,100 (30.4%) | 212 (47.0%) |  |
| Anti-hypertensive medication |  | N | 61,558 | 844 | < 0.001 |
|  | No | n (%) | 45,916 (74.6%) | 483 (65.6%) |  |
|  | Yes | n (%) | 15,642 (25.4%) | 253 (34.4%) |  |
| Systolic blood pressure |  | N | 63,243 | 844 | < 0.001 |
|  |  | mmHg | 135.0 [123.0, 148.0] | 130.0 [118.0, 140.0] |  |
| Diastolic blood pressure |  | N | 63,243 | 844 | < 0.001 |
|  |  | mmHg | 81.0 [74.0, 88.0] | 76.0 [70.0, 81.5] |  |
| Type 2 diabetes |  | N | 62,302 | 844 | < 0.001 |
|  | No | n (%) | 59,610 (95.7%) | 403 (89.2%) |  |
|  | Yes | n (%) | 2,692 (4.3%) | 49 (10.8%) |  |
| Diabetic medication |  | N | 61,558 | 844 | < 0.001 |
|  | No | n (%) | 60,943 (99.0%) | 674 (91.6%) |  |
|  | Yes | n (%) | 615 (1.0%) | 62 (8.4%) |  |
| HbA1c |  | N | 57,908 | – | – |
|  |  | mmol/mol | 34.6 [32.2, 37.0] | – | – |
| Glucose |  | N | 37,223 | – | – |
|  |  | mmol/L | 3.6 [3.1, 4.0] | – | – |
| Lipid-lowering medication |  | N | 61,558 | 844 | < 0.001 |
|  | No | n (%) | 45,761 (74.3%) | 394 (53.5%) |  |
|  | Yes | n (%) | 15,797 (25.7%) | 342 (46.5%) |  |
| HDL |  | N | 36,634 | – | – |
|  |  | mmol/L | 1.3 [1.1, 1.6] | – | – |
| LDL |  | N | 36,628 | – | – |
|  |  | mmol/L | 1.7 [1.5, 2.0] | – | – |
| Triglycerides |  | N | 36,831 | – | – |
|  |  | mmol/L | 1.2 [0.8, 1.6] | – | – |
| White blood cell count |  | N | 59,457 | – | – |
|  |  | 10<sup>9</sup>/L | 6.4 [5.4, 7.5] | – | – |
| Creatinine |  | N | 36,409 | – | – |
|  |  | μmol/L | 66.1 [58.0, 75.2] | – | – |

a. For continuous variables, Median [IQR] are provided and between-group differences were compared using Mann-Whitney U test.
b. For each variable, the first row provides the number of non-missing observations available.
c. For categorical variables, counts of participants (%) are provided and between-group differences were compared using Chi-squared test.
d. Categories with small counts were suppressed as < 10 to reduce disclosure risk.
e. Unable to perform Chi-squared test due to small observation counts.

### 3.2 Identification of location patterns

We identified five reproducible WMH location patterns in the internal dataset (selected by subKMeans and optimal *k* = 5, comparison in eFigure 6). These location patterns are summarised using bullseye representations of the centroids and median regional WMH volumes (Figure 1). Representative examples are provided. **Clusters 1-3** were characterised by lower WMH burdens (median 0.9mL, 1.5mL, and 1.9mL respectively). Cluster 1 showed a higher proportion of WMH in the basal ganglia-internal capsule-thalamus region, Cluster 2 showed posterior predominance in the occipital lobes, and Cluster 3 showed symmetric periventricular involvement. **Clusters 4** and **5** exhibited higher burdens (median 4.4mL and 6.4mL). Cluster 4 displayed frontal predominance, with lesions extending across deeper white matter layers, towards the most juxtacortical layer. Cluster 5 presented a more widespread frontal and parietal involvement, with lesions concentrated in the three inner layers. These location patterns were qualitatively reproduced in OASIS-3. The proportion of participants aged > 65 years increased monotonically from cluster 1 to 5, and the proportions of females were higher in Cluster 3 and Cluster 4 (eFigure 7).

**Figure 1.**
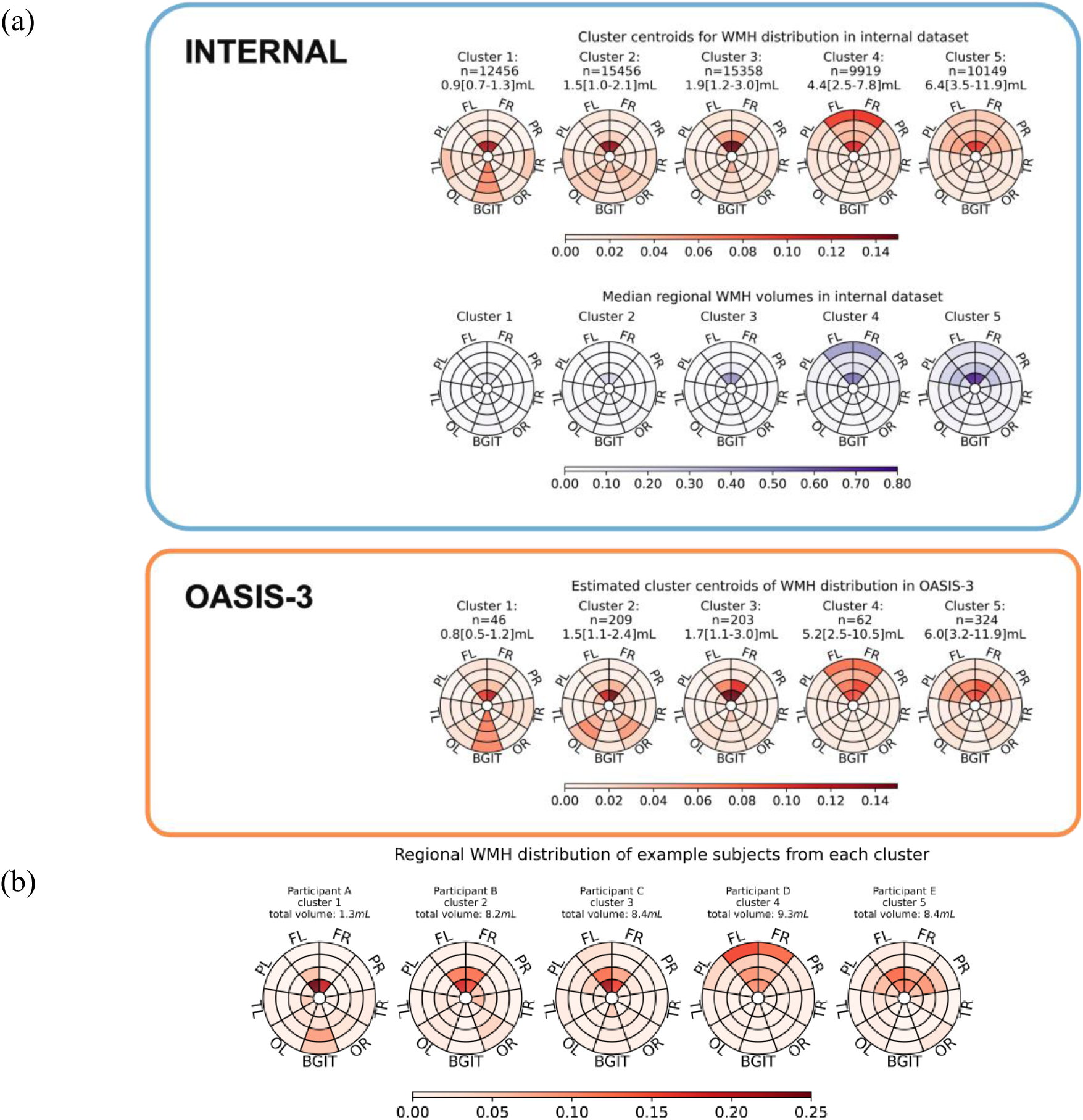
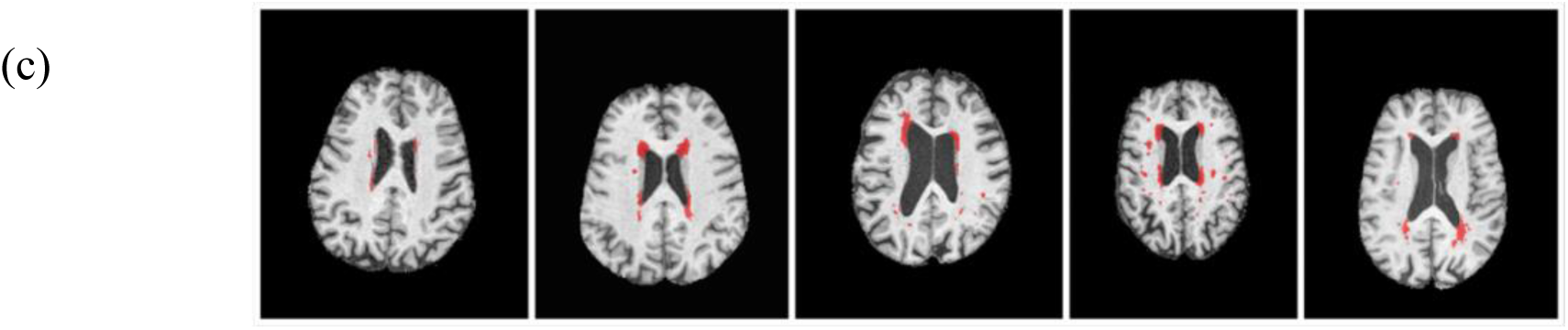
(a) Median WMH regional volumes (mL) in the internal dataset, cluster centroids rescaled to WMH distribution in the internal dataset, and estimated cluster centroids of WMH distribution in OASIS-3, (b) regional WMH distribution in bullseye representation of representative participants and (c) corresponding brain scans in axial view. WMH centroids were rescaled to WMH distribution summing to 1 across 36 regions. The number of participants, median and interquartile range (IQR) of total WMH volumes (mL) in each cluster were labelled for the internal dataset and OASIS-3 respectively. Clusters were ordered by increasing median total WMH volumes from left to right. Median regional WMH volumes were calculated independently for each region within a cluster. The regional values visualised in the bullseye plots do not sum to the median total WMH volume labelled.

#### Sensitivity analyses across age and WMH burden

Sensitivity analyses examined WMH location patterns after stratifying the dataset by baseline WMH volume or age (Figure 2). In the 0-4mL subgroup (n=47,776), five patterns were identified, comprising four low-burden periventricular-focused patterns and one frontal-dominant pattern similar to Cluster 4 in the full dataset. In the 4-8mL subgroup (n=8,634), four patterns corresponding to Clusters 2-5 of the full dataset were observed. In the 67-72 year age group (n=14,745), three location patterns were identified, including two low-burden periventricular patterns and one intermediate pattern between Clusters 4 and 5 of the full dataset.

**Figure 2.**
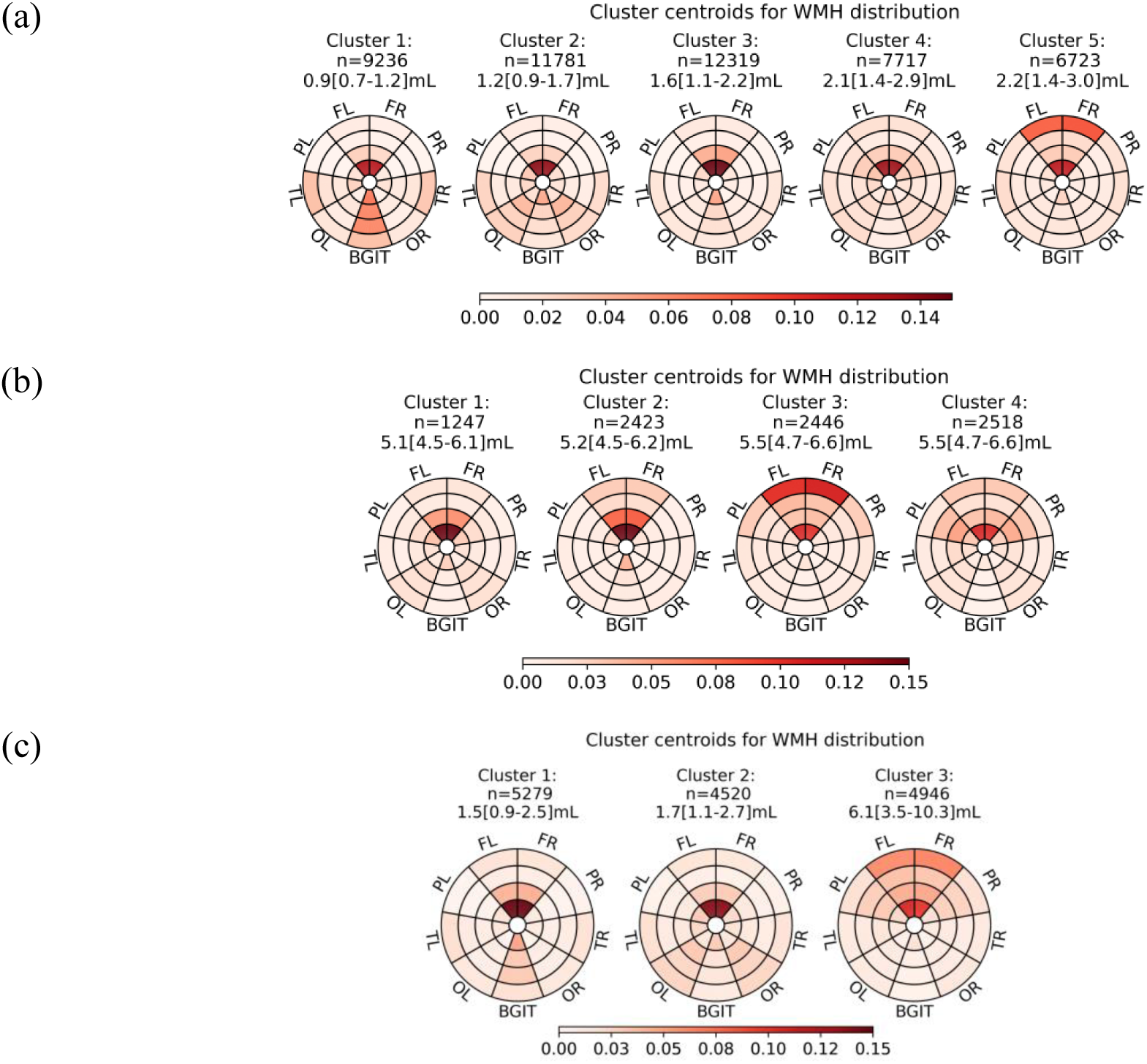
Cluster centroids rescaled to WMH distribution. Clustering repeated in subgroups of (a) 0-4mL total WMH volumes, (b) 4-8mL total WMH volumes, and (c) 67-72 years age group at baseline. Data were rescaled to WMH distribution summing to 1 across 36 regions. The number of participants, median and IQR of total WMH volumes (mL) in each cluster were labelled. Clusters were ordered by increasing median total WMH volumes from left to right.

#### Within-participant shifts between WMH location patterns

Over an 18-30 month follow-up interval (n=3,772), most participants retained the same WMH location pattern (71.5%) (eTable 8). Transitions occurred predominantly within burden strata (Figure 3). Transitions between strata were uncommon and asymmetric. Participants who remained in their baseline cluster were consistently closer to their baseline centroids than those who transitioned, particularly in Clusters 3-5. Individuals who transitioned were already further from their centroids at baseline than those who did not (average distance 0.625 versus 0.580), and larger baseline distances were observed for cross-stratum transitions. No clear pattern was observed between stability and switching behaviour.

**Figure 3.**
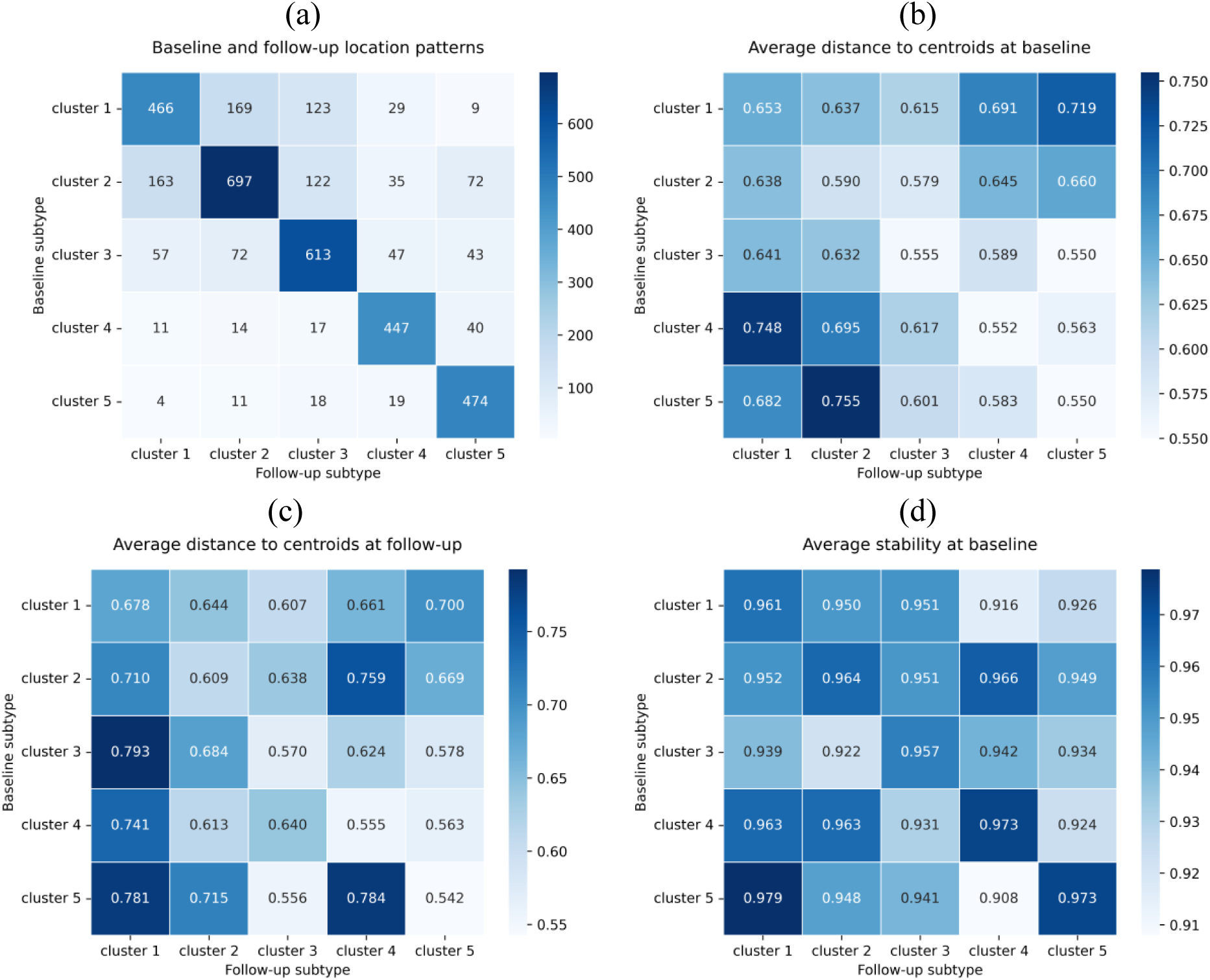
Longitudinal changes in location patterns and cluster stability. Among individuals with 18-30 months follow-up interval, (a) within-participant transitions in location patterns from baseline to follow-up, (b) average distance to centroids at baseline, (c) average distance to centroids at follow-up, and (d) average stability at baseline.

### 3.3 Cross-sectional association between WMH location patterns and risk factors

*Figure 4* shows the cross-sectional associations between 19 non-imaging risk factors and proximity to the centroids of five WMH location patterns, obtained from multiple linear regressions adjusted for age, sex and total WMH volume in the internal data. A positive association indicates that individuals in that cluster were independently associated with higher values in a continuous risk factor or higher odds of belonging to a given category relative to the reference, and vice versa. Full results are provided in *eTable 9* and *eTable 10*.

**Figure 4.**
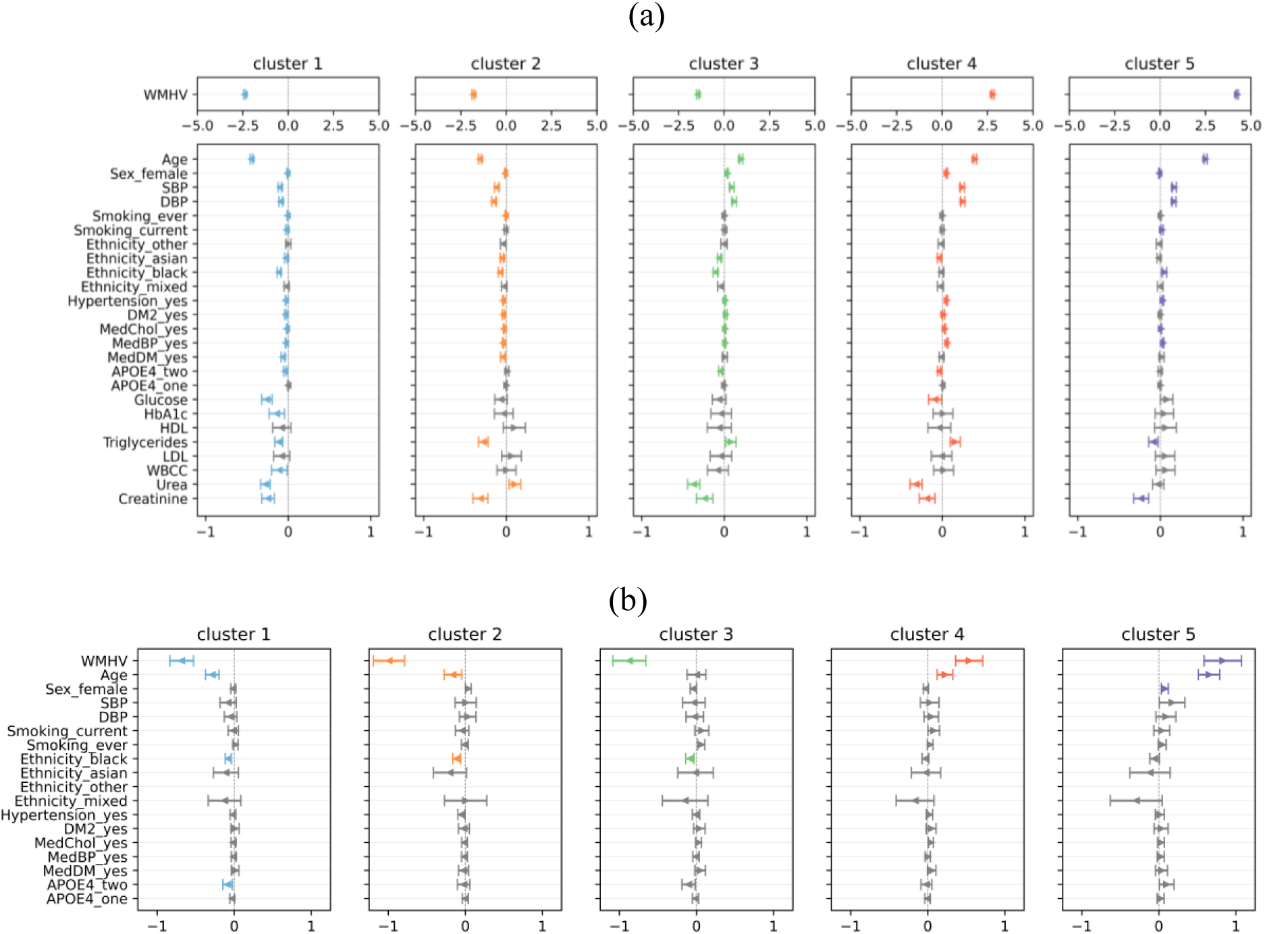
Cross-sectional associations between risk factors and proximity to centroids of 5 clusters in (a) the internal dataset and (b) OASIS-3, estimated using multivariable linear regression adjusted by age, sex and total WMH volume. Each row corresponds to one risk factor or one category. Coloured arrows indicate significant associations (BH-corrected *p* < 0.05), colours are cluster-specific, grey arrows indicate non-significance. Standardised effect sizes (*β*) are represented by their distance from the centre, and the width of error bars indicates the 95% confidence interval (95% CI). The number of participants from Other ethnicity group was insufficient in OASIS-3. Abbreviations: WMHV=total WMH volume, SBP/DBP=systolic/diastolic blood pressure, DM2=type 2 diabetes, MedChol=lipid-lowering medication, MedBP=anti-hypertensive medication, MedDM=diabetic medication, HDL=high-density lipoprotein cholesterol, LDL=low-density lipoprotein cholesterol, WBCC=white blood cell count. Categorical variables and reference categories: sex (male), smoking (never), ethnicity (white), diagnoses of medication conditions or use of medications (no), number of APOE4 alleles (0).

Across all five spatial WMH clusters, age and total WMH volume were strongly associated with centroid proximity. Older age was associated with being closer to Clusters 3-5 (Cluster 3: Effect size *β* = 0.203 [95% CI: 0.183, 0.223], Cluster 4: *β* = 0.393 [0.374, 0.413], Cluster 5: *β* = 0.545 [0.527, 0.564]) and further from Clusters 1-2 (Cluster 1: *β* = -0.446 [-0.462, -0.431], Cluster 2: *β* = -0.320 [-0.338, -0.302]). Higher total WMH volume was associated with greater proximity to Clusters 4-5 (Cluster 4: *β* = 2.792 [2.731, 2.854], Cluster 5: *β* = 4.243 [4.184, 4.301]) and reduced proximity from Clusters 1-3 (Cluster 1: *β* = -2.404 [-2.453, -2.356], Cluster 2: *β* = -1.816 [-1.875, -1.758], Cluster 3: *β* = -1.461 [-1.525, -1.397]). Higher systolic, diastolic blood pressure, hypertension, Type 2 diabetes were each independently associated with greater proximity to Clusters 1 and 2 and reduced proximity to Clusters 3-5 (Type 2 diabetes only showed significant associations with Clusters 4-5).

Lifestyle and demographic factors further differentiated clusters. Current smokers were further from Cluster 1 (*β* = -0.021 [-0.033, -0.009]) and closer to Cluster 5 (*β* = 0.021 [0.006, 0.035]). Ever smokers were further from Clusters 1 and 2. Female sex was associated with being closer to Cluster 3 (*β* = 0.040 [0.035, 0.046]) and to Cluster 4 (*β* = 0.054 [0.049, 0.060]). Asian ethnicity was consistently associated with being further from Clusters 1-4, with no association reaching statistical significance for Cluster 5. Black ethnicity showed reduced proximity to Clusters 1-3 yet greater proximity to Cluster 5.

Patients having type 2 diabetes were further from Clusters 1 and 2 and closer to Clusters 3-4. The effects of APOE4 were small, with two copies associated with being further from Clusters 1, 3, and 4. Medication use was predominantly associated with higher-burden patterns. Among metabolic, inflammatory and renal markers, Cluster 1 displayed the greatest number of associations, characterised by lower levels of glucose, triglycerides, and creatinine. Triglycerides differentiated lower- and higher-burden patterns, with reduced proximity to Clusters 1-2 (*e.g.* Cluster 2: *β* = -0.114 [-0.161, -0.067]) and increased proximity to Clusters 3-4 (*e.g.* Cluster 4: *β* = 0.157 [0.098, 0.217]), except for Cluster 5 (*β* = -0.082 [-0.139, -0.025]). WBCC, LDL, and HDL showed minimal or no significant associations. Higher creatinine level was consistently associated with reduced proximity to all clusters.

While sharing similar directionality, effect estimates in OASIS-3 were generally attenuated compared with the internal dataset.

### 3.4 Comparing risk factors and location patterns between WMH progressors and non-progressors

Distribution of risk factors significantly different between progressors and non-progressors is presented in Figure 5 and eTable 11 (internal total: n=5,274, progressors: n =1,495, non-progressors: n=3,779). Compared to non-progressors, progressors had larger baseline WMH volume, were older (p < 0.001) with higher blood pressures (SBP and DBP: p < 0.001). Although differences were often marginal, progressors had higher HbA1c levels and lower HDL, LDL, and WBCC (all p < 0.001). More progressors took lipid-lowering medications (p=0.006) or carried at least one APOE *ε*4 allele. Progressors also tended to be closer to Clusters 4 and 5, and further from Clusters 1 and 2 (all p < 0.001). In OASIS-3 (n=185, progressors: n=94, non-progressors: n=88), only age and participants’ distance to the centroid of cluster 1 and 5 were different (both p < 0.001).

**Figure 5.**
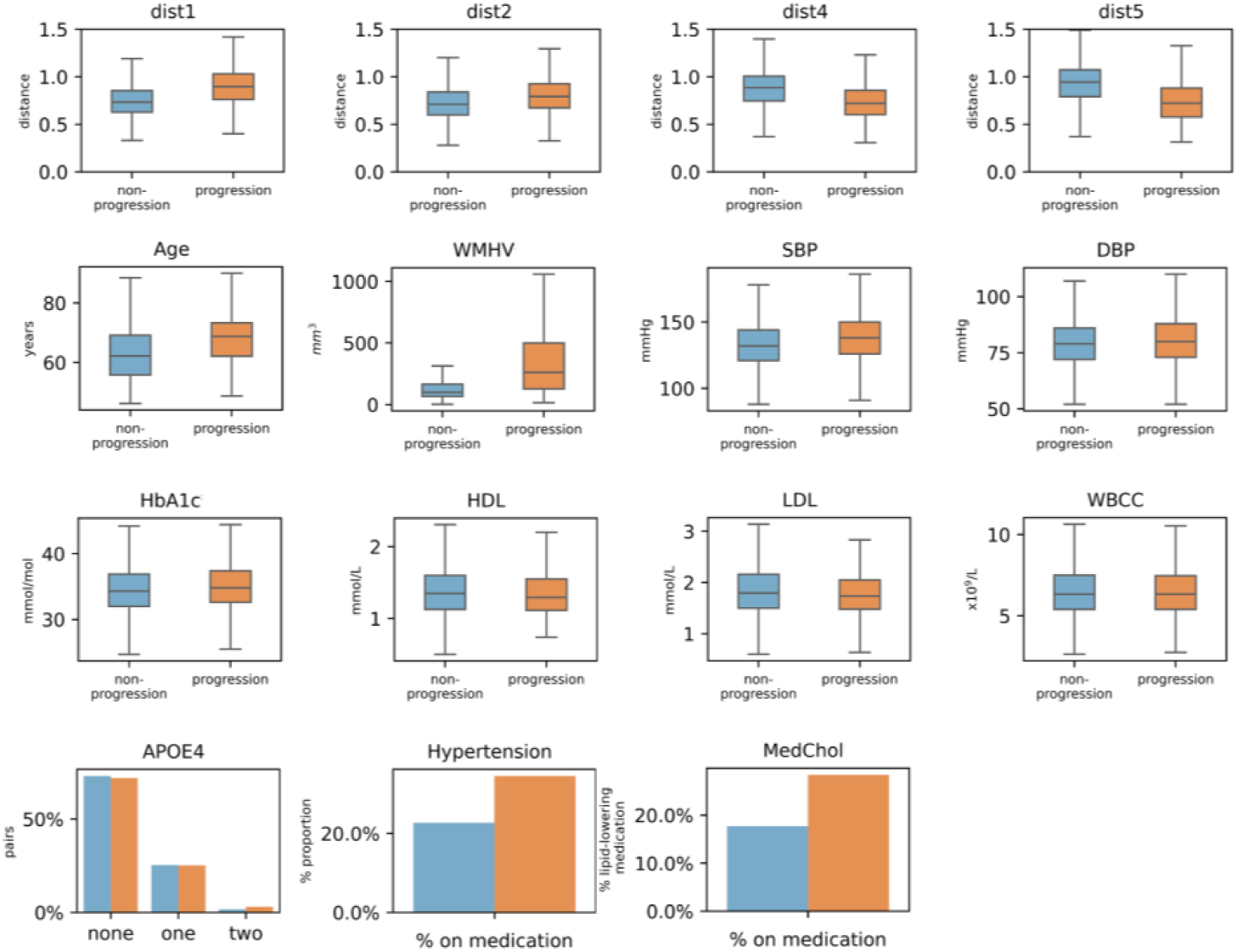
Distribution of Euclidean centroids, risk factors and total baseline WMH volume between progression and non-progression groups in the internal dataset. The two groups were compared after removing the effect of age and sex, where applicable. P-values were BH-corrected. Only significantly different variables are presented. Abbreviations: dist1-5: distance to the centroid of clusters 1-5, WMHV=total baseline WMH volume, SBP/DBP=systolic/diastolic blood pressure, WBCC=white blood cell count, MedChol=lipid-lowering medication.

### 3.5 Longitudinal prediction of WMH progression

The internal training, testing set and external OASIS-3 set comprised 1,188 (28.2%), 307 (29.1%) and 88 (51.6%) progressors respectively. Model performance is reported in Figure 6 and Section e2.8.

**Figure 6.**
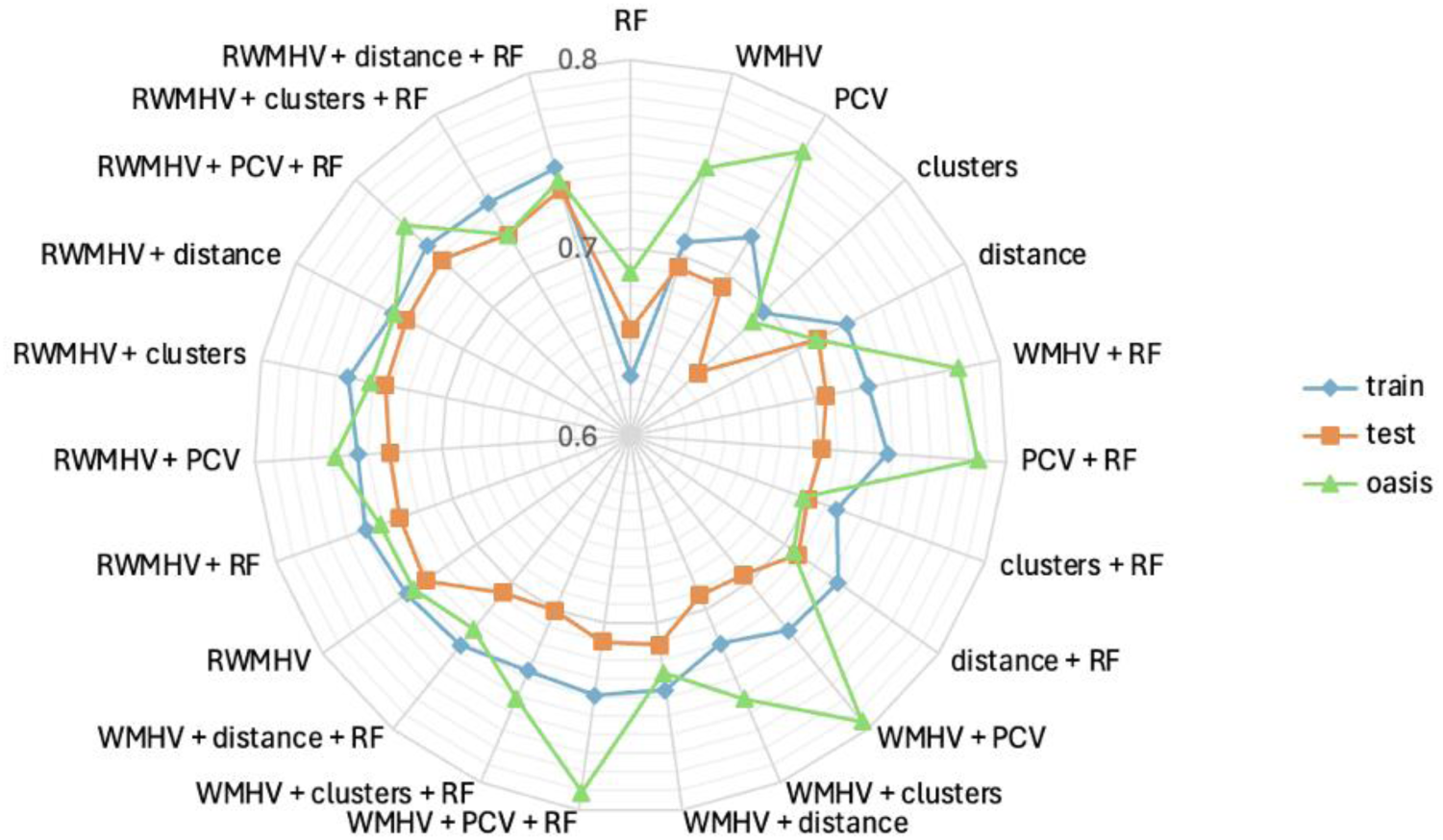
Mean balanced accuracy of different predictor combinations in training, test set and OASIS-3 for WMH progression. Abbreviations: clusters=location patterns of WMH, distance=Euclidean distance to five cluster centroids, RF=age + sex + systolic blood pressure, WMHV=total baseline WMH volume, RWMHV=36 regional baseline WMH volume, PCV=principal components of RWMHV.

Models incorporating regional WMH volumes (RWMHV, below refers to models by abbreviations) achieved the strongest performance in predicting WMH progression. In the test set, WMHV or RWMHV alone reached average balanced accuracies of 0.693 (95% CI: 0.664-0.723) and 0.733 (0.703-0.762) respectively. The best-performing model additionally incorporated principal components of volumes (PCV) and risk factors (RF) (RWMHV + PCV + RF: 0.737, 0.706-0.764), but the improvement over RWMHV-only model was marginal and comparable to adding centroid distances (RWMHV + distance: 0.734, 0.704-0.761) or distance plus RF (RWMHV + distance + RF: 0.736, 0.707-0.762). Notably, adding PCV or discrete cluster assignment to RWMHV did not consistently improve performance and in several cases slightly reduced balanced accuracy. Meanwhile, the inclusion of centroid distances made small but consistent improvements. Distance-only models achieved reasonable performance despite including only five variables (0.712, 0.681-0.742). PCV-only models performed less well (0.693, 0.662-0.722). In OASIS-3, RWMHV-based models also performed consistently well, with average balanced accuracy ranging between 0.734 (RWMHV: 0.670-0.795) and 0.757 (RWMHV + PCV + RF: 0.696-0.815). However, several WMHV- and PCV-augmented models showed substantially higher performance in OASIS-3 than in the internal test set.

## 4 Discussion

### 4.1 Five WMH location patterns

In this multi-cohort study of ageing populations from the UK and USA, we identified five distinct WMH location patterns by clustering individuals based on the relative spatial distribution of WMH across 36 brain regions. The stability selection approach demonstrated the reproducibility of data-driven subtyping. Unlike voxel-level clustering methods requiring registration to a common space [16] or bullseye-based PCA approaches that extract latent spatial components from regional WMH volumes [10], [14], [15], our approach provides discrete and interpretable representations that emphasised WMH distribution independent of total burden.

Although the patterns varied in overall WMH burden, these differences did not explain their distinct spatial profiles. In particular, high-burden clusters varied markedly: WMH extended into deeper white matter regions in Cluster 4, whereas a more superficial and diffuse involvement was observed in Cluster 5. In addition, we were able to stratify individuals with similar overall WMH burden into different location patterns. Sensitivity analyses confirmed that the main findings were stable across strata of age and WMH volume. Longitudinal analyses over 18-30 months found substantial within-participant stability and limited transitions from high-burden to low-burden patterns. The WMH patterns were qualitatively replicated in OASIS-3. Overall, our location patterns were broadly consistent with prior spatial WMH phenotyping studies. As discussed in Section 1, one study identified five voxel-level WMH spatial signatures including periventricular, deep frontal, posterior, and more superficial or juxtacortical signatures [16]. Multiple bullseye-based analyses demonstrated that WMH spatial heterogeneity could be captured by 3-7 principal components that reflected periventricular, deep, and more superficial involvement [10], [14], [15].

### 4.2 Characterising WMH location patterns

Analyses of association with risk factors revealed distinct risk profiles between clusters. Total WMH volume was the strongest factor, with higher-burden patterns aligning most strongly with Clusters 4-5 and low-burden patterns with Clusters 1-2. This was followed by age, where older age aligned with Clusters 3-5 and younger age with Clusters 1-2. Compared with low-burden patterns, higher-burden patterns aligned with less favourable clinical profiles (*e.g.* higher blood pressures, current smoking, diabetes and increased medication use) independent of age, sex and baseline WMH burden, indicating a gradient of disease severity. Females were closer to Clusters 3 and 4. Participants of black ethnicity were closer to Clusters 5 but further from Cluster 1-3. Asian ethnicity did not align with any location patterns, suggesting they may not generalise well to Asian populations. Cluster 1 displayed the most favourable profile from blood-derived markers, while most lipid or inflammatory markers were not differential between WMH patterns. In OASIS-3, the directions of associations were generally replicated, although effect sizes were generally smaller and statistical significance was attenuated due to substantially reduced sample size and statistical power.

The dominant role of age agreed with previous studies [10], [14], [15], [16]. Pattern- or component- specific findings on blood pressure, hypertension and diabetes were also reported [10], [16]. Our observations on the statistically significant but small effects of APOE4 alleles is compatible with previous null findings [10], [16] given their limited sample size (less than 3,000 participants) [10]. Associations between posterior deep WMH and two APOE4 alleles were previously reported in UKB [49]. Several associations differed across studies. We observed smoking and sex-related pattern alignment, which was again consistent with one relevant study [49]. However, other studies reported mixed or null associations [10], [14], [15]. Ethnicity-related variation was evident in our data, while the only relevant study that included ethnicity did not find significant differences in associations between Blacks and Whites [16]. Finally, our systematic evaluation of metabolic, inflammatory, and renal blood markers extended prior spatial phenotyping work, which focused primarily on demographic, vascular and genetic factors (except for renal CIRS scores [15]).

### 4.3 Implications for longitudinal prognosis

We found WMH progressors aligned more with high-burden clusters and tended to have adverse vascular risk profiles, including older age, higher baseline WMH burden, and elevated blood pressure.

Consistent with prior work, baseline WMH volume was as a strong predictor of WMH progression [33], [34]. Models including regional WMH volumes consistently outperformed other candidates, highlighting the prognostic value of spatially defined WMH burden. Although the best-performing model additionally incorporated principal components derived from regional WMH volumes, the improvement over regional volumes alone was marginal. However, none of the contributions from dimensionality-reduced representations (principal component scores, discrete cluster assignments, or distance to centroids) were statistically significant. The differences in absolute performance observed in OASIS-3 may reflect cohort composition or follow-up characteristics.

### 4.4 Limitations

This study has several limitations. First, data were drawn from multiple cohorts without harmonisation of imaging protocols, introducing potential scanner- and site-related variability that may influence clustering and prediction. Second, the dominant contribution of the comparatively younger and healthier UKB cohort may bias spatial WMH subtypes toward patterns less representative of clinical populations. This may explain why our findings aligned closely with a UKB-based study on smoking, diabetes, and APOE4 [49]. Asian and Black participants were under-represented in our data, which may limit the generalisability of our findings to more ethnically diverse populations. Ethnicity was recorded only at a broad level that combined populations with heterogeneous origins and cultures. Fourth, stroke status was not considered, despite its strong influence on WMH burden and progression. Post-stroke WMH may transiently regress due to oedema resolution [37], and failure to account for this may affect the prediction of longitudinal trajectories [50]. Other unmeasured confounders, such as socioeconomic status and education, may also influence WMH distribution and evolution. Furthermore, while distinct spatial subtypes of WMH were identified and associated with certain risk profiles, their underlying pathophysiological basis was not discussed. Sixth, although baseline location patterns demonstrated significant associations with WMH progression and reasonable predictive value, they did not outperform regional WMH volumes in both prediction tasks. In the OASIS-3 cohort, longitudinal scans were available for only a smaller and older subset, limiting statistical power to detect differences. Further, OASIS-3 had a more balanced progressor-to-non-progressor ratio (internal: 1:3, OASIS-3: 1:1), which raised a question about how much the fixed thresholds were transferable between studies [46]. Finally, scans from only one follow-up were available and limited our ability to capture non-linear WMH changes over a longer period. Future studies with a broader temporal resolution could help improve understanding of WMH dynamics.

## 4.5 Conclusion

In summary, our study presented a robust data-driven framework for identifying and characterising location patterns of WMH across different studies. Such patterns showed substantial temporal stability and supported their interpretation as meaningful phenotypes of CSVD by demonstrating their distinct associations with demographic, cardiovascular, metabolic, renal, inflammatory, and genetic risk factors. Regional WMH burden remained the best candidate in longitudinal prediction, while spatial pattern information provided complementary insights into WMH heterogeneity beyond global lesion volume. These findings provided a basis for future work in image-based prediction of WMH progression. Overall, our study highlighted the role of spatial phenotyping in characterising WMH heterogeneity beyond lesion volume, while emphasising the importance of regional WMH burden for predicting progression.

## Data availability

Data used in this study were obtained from ADNI3, NSHD, SABRE and UKB databases. ADNI3 data can be requested and downloaded from https://adni.loni.usc.edu/. The data are publicly available to qualified scientific investigators upon submission of an online application and formal acceptance of the ADNI Data Use Agreement. As per the terms of this agreement, the authors cannot redistribute ADNI data. Access to NSHD data is governed by the NSHD and is available to bona fide researchers via application through the NSHD Skylark data sharing platform (https://skylark.ucl.ac.uk/NSHD/). SABRE data is managed by University College London (UCL). Access to SABRE data for bona fide research purposes can be requested by submitting a research proposal to the study’s data access committee, and access is subject to a formal Data Sharing Agreement. The UK Biobank data was obtained under Application Number 12113. The UK Biobank resource is available to bona fide researchers for health-related research in the public interest. Access requires registration and an approved application via the Access Management System (https://ams.ukbiobank.ac.uk/), execution of a Material Transfer Agreement, and payment of an access fee. The OASIS-3 dataset is available upon request at https://sites.wustl.edu/oasisbrains/home/oasis-3/.

## Acknowledgement

Data collection and sharing for this project were funded by the Alzheimer’s Disease Neuroimaging Initiative (ADNI) (National Institutes of Health Grant U01 AG024904) and DOD ADNI (Department of Defense award number W81XWH-12-2-0012). ADNI is funded by the National Institute on Aging, the National Institute of Biomedical Imaging and Bioengineering, and through generous contributions from the following: AbbVie, Alzheimer’s Association; Alzheimer’s Drug Discovery Foundation; Araclon Biotech; BioClinica, Inc.; Biogen; Bristol-Myers Squibb Company; CereSpir, Inc.; Cogstate; Eisai Inc.; Elan Pharmaceuticals, Inc.; Eli Lilly and Company; EuroImmun; F. Hoffmann-La Roche Ltd and its affiliated company Genentech, Inc.; Fujirebio; GE Healthcare; IXICO Ltd.; Janssen Alzheimer Immunotherapy Research & Development, LLC.; Johnson & Johnson Pharmaceutical Research & Development LLC.; Lumosity; Lundbeck; Merck & Co., Inc.; Meso Scale Diagnostics, LLC.; NeuroRx Research; Neurotrack Technologies; Novartis Pharmaceuticals Corporation; Pfizer Inc.; Piramal Imaging; Servier; Takeda Pharmaceutical Company; and Transition Therapeutics. The Canadian Institutes of Health Research is providing funds to support ADNI clinical sites in Canada. Private sector contributions are facilitated by the Foundation for the National Institutes of Health (www.fnih.org). The grantee organization is the Northern California Institute for Research and Education, and the study is coordinated by the Alzheimer’s Therapeutic Research Institute at the University of Southern California. ADNI data are disseminated by the Laboratory for Neuro Imaging at the University of Southern California.

We are very grateful to the NSHD study members who helped design and conduct the study and to the participants for their contributions to Insight 46 and for their commitments to research over the last seven decades, and to the past and current members of the Insight 46 study team.

We thank all the SABRE participants who took part in the study, and past and present members of the SABRE team who helped to collect the data.

This research has been conducted using the UK Biobank Resource under Application Number 12113. Please see https://www.ukbiobank.ac.uk/ for more information.

Data were provided in part by OASIS-3: Longitudinal Multimodal Neuroimaging: Principal Investigators: T. Benzinger, D. Marcus, J. Morris; NIH P50 AG00561, P30 NS09857781, P01 AG026276, P01 AG003991, R01 AG043434, UL1 TR000448, R01 EB009352. AV-45 doses were provided by Avid Radiopharmaceuticals, a wholly owned subsidiary of Eli Lilly.

## Footnotes

1 Data used in the preparation of this article were obtained from the Alzheimer’s Disease Neuroimaging Initiative (ADNI) database (adni.loni.usc.edu). ADNI was launched in 2003 as a public-private partnership, led by Principal Investigator Michael W. Weiner, MD. The primary goal of ADNI has been to test whether serial magnetic resonance imaging (MRI), positron emission tomography (PET), other biological markers, and clinical and neuropsychological assessment can be combined to measure the progression of mild cognitive impairment (MCI) and early Alzheimer’s disease (AD).

2 From https://github.com/KCL-BMEIS/NiftySeg

3 The additional thalamus and infratentorial region were combined into basal ganglia in subsequent processing

4 Ethnicity was not available in Insight46. “Mixed” group was not available in SABRE, “Other” group was not available in OASIS-3.

5 For categorical variables, the first categories declared were the reference categories where applicable.

6 Implemented using scikit-learn [39].

7 Implemented using scikit-learn [39].

8 Implemented using scikit-learn [39].

9 Performance evaluation implemented using scikit-learn [39].

