## Supplementary materials for "Location patterns and longitudinal progression of white matter hyperintensities"

#### e1 Methods

##### e1.1 Datasets

The multi-cohort dataset included ADNI3 scans collected between 2017 and 2022, Insight46 scans collected between 2015 and 2019, SABRE scans collected between 2014 and 2019, UKB scans since 2014, and OASIS-3 collected over the course of 30 years.

**ADNI3** ADNI3 is a multi-centre, natural history, prospective study based in North America [18], [19], [20]. Data collection for ADNI3 began in 2016, and was expected to conclude by the end of 2024. Participants were aged 55-90 at baseline, with no significant neurological disease other than Alzheimer's disease. In addition to MRI scans, a comprehensive set of demographic information (*e.g.* age, sex), medical history (*e.g.* systolic and diastolic blood pressures), neuropsychology tests (*e.g.* ADAS11/13, MMSE), positron emission tomography (PET) and genotype data (*e.g.* APOE4 genotype) are also available.

**Insight46** Insight46 is the prospective neuroimaging substudy of the MRC National Survey for Health and Disease (NSHD) birth cohort [21], [22]. It drew a subset of 502 participants from this cohort of individuals all born in the same week of March 1946. Baseline neuroimaging data were collected between 2015 and 2017, when participants were aged 69-71, and follow-up data were collected between 2018 and 2020, when participants were aged 73-75. The study collected clinical, neuropsychological, brain MRI scans, PET amyloid imaging, blood, urine and CSF biomarkers.

**SABRE** The SABRE (Southall and Brent Revisited) cohort study is a longitudinal project based in West London [23]. It originally focused on cardiovascular disease and diabetes in people of White British, first-generation migrants of South Asian or African Caribbean heritage. Baseline data were collected between 1988 and 1991 (when participants were aged between 40 and 69). At the third wave of data collection (2014-2019), neuroimaging data were acquired for 772 participants and their spouses on a 3T Philips Achieva Scanner.

**UKB** UK Biobank recruited approximately 500,000 participants between 2006 and 2010 [24]. Data collected include demographic, lifestyle, clinical, biochemical, genetic and imaging measures, with linkage to electronic health records. A subset of approximately 67,000 participants underwent brain MRI scans [25], with follow-ups available for some participants.

**Validation dataset: OASIS-3** OASIS-3 is a longitudinal neuroimaging, clinical, cognitive, and biomarker dataset that retrospectively collected data from 1378 participants in the United States aged between 42 and 95 [26]. It includes participants who were cognitively normal as well as those at various stages of cognitive impairment and dementia. Data collection was conducted at a single site at the Knight Alzheimer Disease Research Center at Washington University in St. Louis, Missouri.

##### e1.2 Quality control

Following segmentation, we first conducted a visual inspection of WMH masks from ten ADNI3 scans. This was followed by further inspection of scans from both UKB and ADNI3 datasets, focusing on suspected anomalies - specifically, those with either a high volume of WMH lesions (over 80mL) or an absence of WMH lesions. Any incorrect or erroneous records were either re-processed (ADNI3: n=12 at baseline, UKB: 4 at follow-up) or excluded (ADNI3: n=1 at baseline, n=2 at follow-up, UKB: n=16 at follow-up). Flowcharts for participants of each cohort are provided in e1.6.

#### e1.3 MRI protocol

*eTable 1 MRI acquisition parameters across cohorts.*

| Cohort | Scanner vendor / model | Site | Field strength | Sequence | TR (ms) | TE (ms) | TI (ms) | Voxel size ($mm^3$) |
|---|---|---|---|---|---|---|---|---|
| **ADNI3** | GE, Siemens, Philips | Multi | 3T | T1 | 2300 | min full echo | 900 | 1×1×1 |
| | | | | T2-FLAIR | 4800 | 119 | 1650 | 1.2×1×1 |
| **Insight46** | Biograph mMR 3T PET/MRI | Single | 3T | T1 | 2000 | 2.92 | 870 | 1.1×1.1×1.1 |
| | | | | T2-FLAIR | 5000 | 402 | 1800 | 1.1×1.1×1.1 |
| **SABRE** | Philips Achieva | Single | 3T | T1 | 6.9 | 3.1 | - | 1×1×1 |
| | | | | T2-FLAIR | 4800 | 125 | 1650 | 1×1×1 |
| **OASIS-3** | Siemens (various models) | Single | 3T, some 1.5T | T1 | 2400 | 3.16 | 1000 | 1×1×1* |
| | | | | T2-FLAIR | 900 | 91 | 2500 | 1×1×1* |
| **UK Biobank** | Siemens Skyra | Multi | 3T | T1 | 2000 | - | 880 | 1×1×1 |
| | | | | T2-FLAIR | 5000 | - | 1800 | 1.05×1×1 |

** Note: Lower resolutions also present.*

#### e1.4 Availability of variables

*eTable 2 Availability of non-imaging variables across cohorts. ✓indicates that the variable is available.*

| VARIABLE | ADNI-3 | INSIGHT46 | SABRE | UKB | OASIS-3 |
|---|---|---|---|---|---|
| Age | ✓ | ✓ | ✓ | ✓ | ✓ |
| Sex | ✓ | ✓ | ✓ | ✓ | ✓ |
| Ethnicity | ✓ | | ✓ | ✓ | ✓ |
| Total WMH volume | ✓ | ✓ | ✓ | ✓ | ✓ |
| APOE4 genotype | ✓ | ✓ | ✓ | ✓ | ✓ |
| Smoking status | | ✓ | ✓ | ✓ | ✓ |
| Hypertension | ✓ | ✓ | ✓ | ✓ | ✓ |
| Anti-hypertensive medication | | ✓ | ✓ | ✓ | ✓ |
| Systolic blood pressure | ✓ | ✓ | ✓ | ✓ | ✓ |
| Diastolic blood pressure | ✓ | ✓ | ✓ | ✓ | ✓ |
| Type 2 diabetes | | ✓ | ✓ | ✓ | ✓ |
| Diabetic medication | | ✓ | ✓ | ✓ | ✓ |
| HbA1c | | ✓ | ✓ | ✓ | |
| Glucose | ✓ | ✓ | ✓ | ✓ | |
| Lipid-lowering medication | | ✓ | ✓ | ✓ | ✓ |
| LDL cholesterol | | ✓ | ✓ | ✓ | |
| HDL cholesterol | | ✓ | ✓ | ✓ | |
| Triglycerides | ✓ | ✓ | ✓ | ✓ | |
| White blood cell count | | ✓ | ✓ | ✓ | |
| Creatinine | ✓ | ✓ | ✓ | ✓ | |

#### e1.5 Definition of variables

##### Ethnicity

*eTable 3 Definition of ethnicity across cohorts.*

| HARMONISED LABEL | ADNI3 | INSIGHT46 | SABRE | UKB | OASIS-3 |
|---|---|---|---|---|---|
| White | White | Not available | European | 1 | White |
| Black | Black | Not available | African Caribbean | 4 | Black |
| Asian (incl. Chinese) | Asian | Not available | South Asian | 3, 5 | Asian |
| Mixed | More than one | Not available | N/A | 2 | more than one |
| Other | Unknown | Not available | Other | 6 | Unknown |

**Smoking status**

*eTable 4 Definition of smoking status across cohorts.*

| HARMONISED LABEL | ADNI3 | INSIGHT46 | SABRE | UKB | OASIS-3 |
|---|---|---|---|---|---|
| Never smoker | Not available | Never | Never | Never | |
| Former smoker | Not available | Ex | Ex | Ever | (processed) |
| Current smoker | Not available | Current | Current | Current | |

Note: ADNI3 does not have smoking status recorded near the visit used for analysis. OASIS-3 smoking status was processed from other variables.

In OASIS-3, smoking status was derived using a hierarchical classification to prioritise recent behaviour. Participants were classified as current smokers if TOBAC30=1, indicating cigarette use in the past 30 days, with this category taking priority over all others. Ex-smokers were defined as those with TOBAC30=0 and evidence of past smoking, determined by at least one of the following: TOBAC100=1 (more than 100 cigarettes smoked in lifetime), SMOKYRS> 0 (non-zero years of smoking), valid QUITSMOK data (reported age at quitting), or valid PACKSPER data (reported packs per day). This category required both non-current smoking and a positive smoking history. Never smokers were defined as participants with TOBAC30=0 and TOBAC100=0, and no smoking history. Those who met all of: SMOKYRS null or 0, no quit age (QUITSMOK null), and no reported packs per day (PACKSPER null) were considered to have no smoking history. Participants with missing or insufficient information to meet these criteria were classified as unknown.

**Medication use**

*eTable 5 Data field names used for medication variables across cohorts.*

| HARMONISED LABEL | ADNI3 | INSIGHT46 | SABRE | UKB | OASIS-3 |
|---|---|---|---|---|---|
| Lipid-lowering | n/a | vasc_cholmed_bin_i46p1 | q4_lipidloweringmedsv3 | field 6177 (males) and 6153 (females), field 131286 (date first reported essential primary hypertension), 131294 (date of secondary hypertension) | processed from medname |
| Diabetic | | vasc_bplwrmed_bin_i46p1 | q4_gliptinv3, q4_glitazonev3, q4_glp1v3, q4_insulinv3, q4_metforminv3, q4_sureav3 | | |
| Anti-hypertensive | | vasc_diabeticmed_i46p1 | q4_antihypertensiveany_medsv3, q3_10b_highbpcurrentmedsv3 | | |

#### e1.6 Clustering implementation

The R implementation of the stability estimation method is publicly available at https://github.com/hyu-ub/bootcluster [44]. We translated the relevant code into Python and made several modifications to suit the requirements of the analysis. Key modifications included: 1) Enabled additional clustering methods beyond k-means, as used in the original study, 2) Added outputs of cluster-wise stabilities, in addition to the original global and observational-level stabilities, 3) Incorporated functionality to reuse stability profiles for downstream tasks, and streamline experimental testing.

#### e1.7 5-fold cross-validation in clustering

We performed internal validation to select the optimal k. The dataset was divided into five folds, analogous to a 5-fold cross-validation procedure in classification tasks. For each iteration, stability estimation was performed at the optimal k (as well as the optimal clustering method) on the four training folds and fit to test fold before evaluation. We assumed the clustering on the entire internal dataset to be the ground truth, and map the training and original cluster assignments to each other for comparison.

#### e1.8 Longitudinal prediction

**Uncertainty estimation of prediction models** Training performance was summarised across five cross-validation folds. Balanced accuracy was summarised as the mean and ±2 standard deviations. Test and OASIS-3 performance uncertainty was estimated using 1,000 non-parametric bootstrap iterations, from which the mean and 95% confidence interval (2.5-97.5 percentiles) of balanced accuracy were derived.

### e2 Results

#### e2.1 Participant flowchart

*eFigure 1 Flowchart for ADNI3 participants*

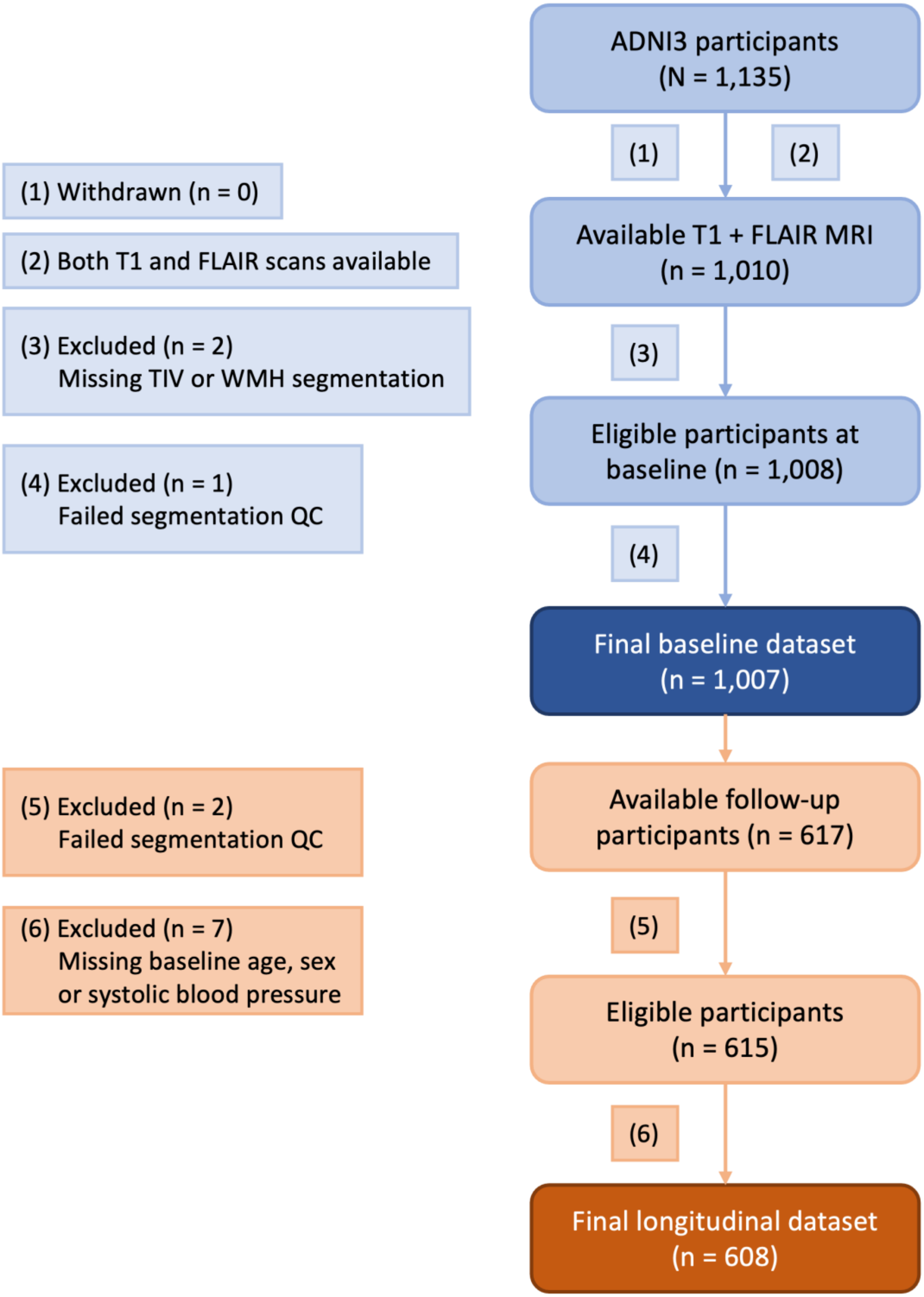

*eFigure 2 Flowchart for Insight46 participants*

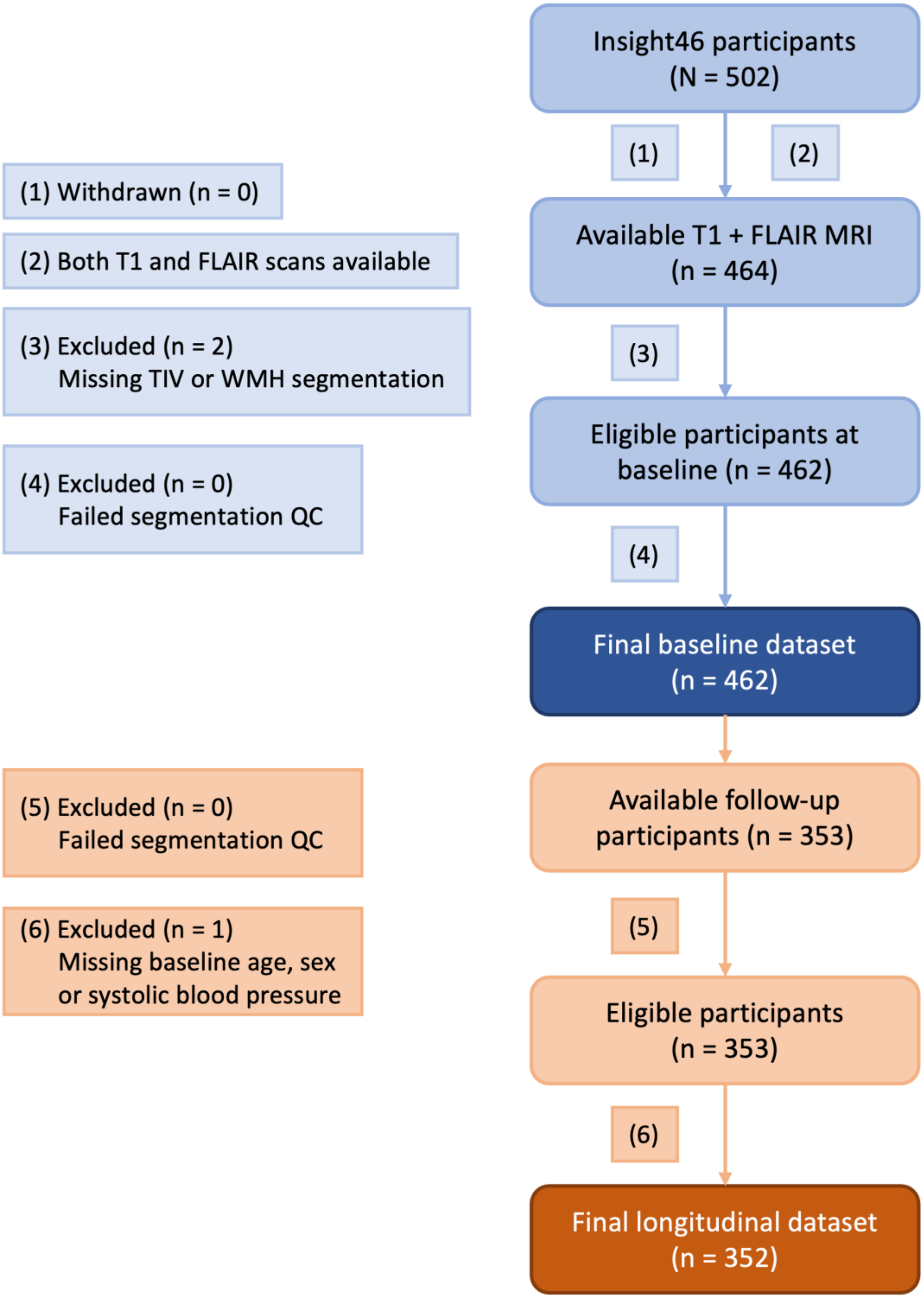

*eFigure 3 Flowchart for SABRE participants*

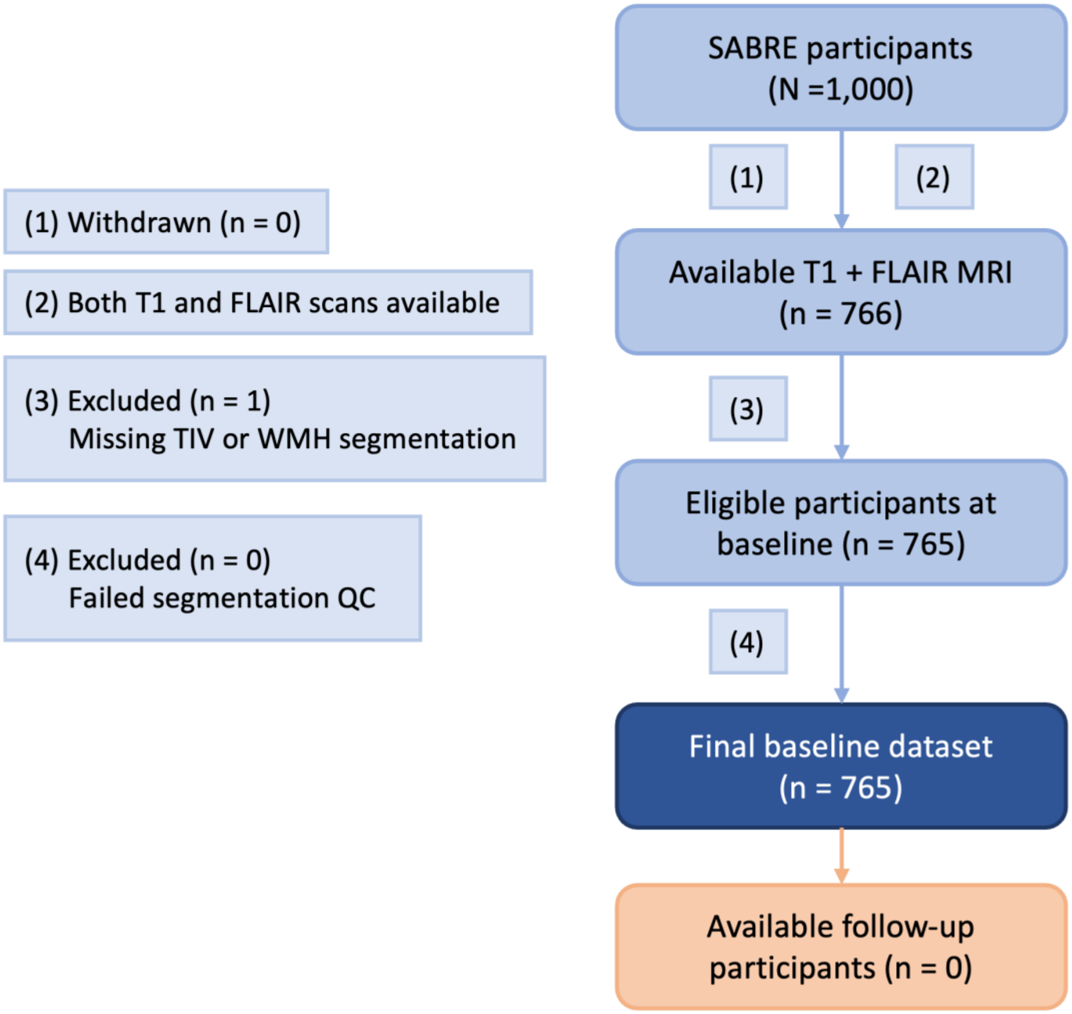

*eFigure 4 Flowchart for UKB participants*

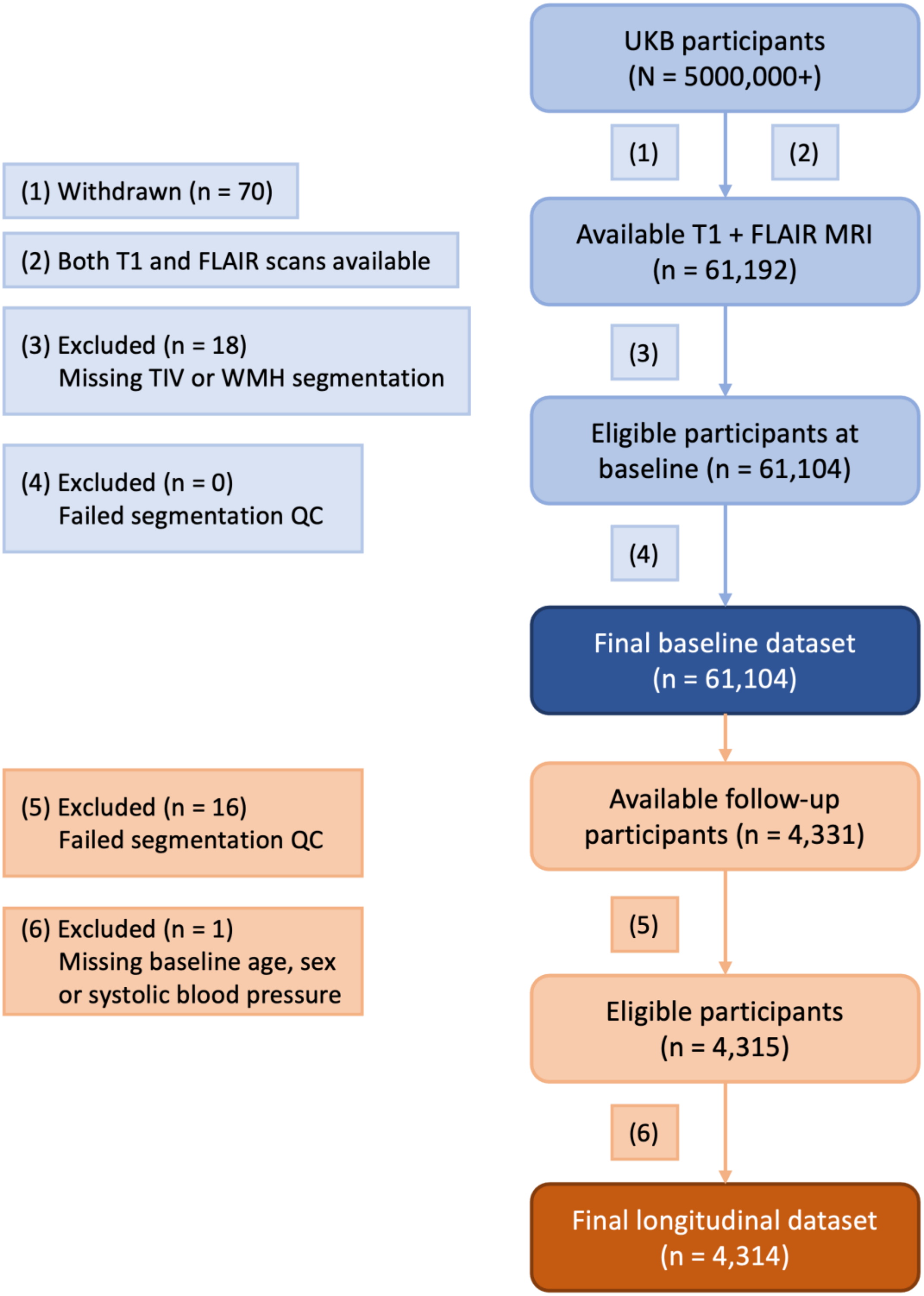

*eFigure 5 Flowchart for OASIS-3 participants*

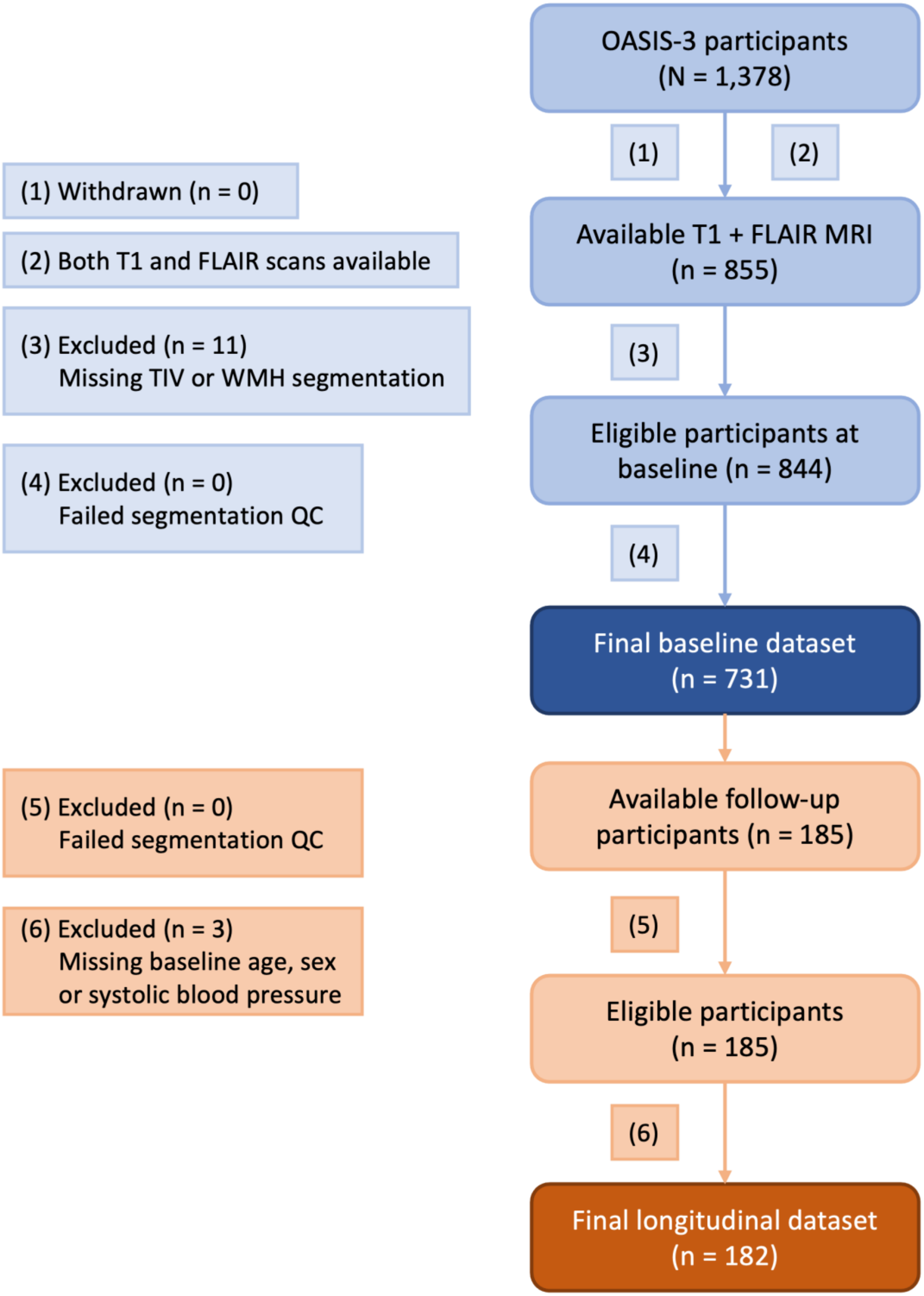

### e2.2 Baseline characteristics by cohort

#### *eTable 6 Baseline characteristics across cohorts.*

| | CATEGORY | UNIT | ADNI-3 (N=1,007) | INSIGHT46 (N=462) | SABRE (N=765) | UKB (N=61,104) | OASIS-3 (N=844) |
|---|---|---|---|---|---|---|---|
| Age[a] | | N[b] | 984 | 462 | 476 | 61,104 | 731 |
| | | years | 72.8 [67.4, 78.6] | 70.7 [70.1, 71.2] | 72.6 [69.2, 76.9] | 65.9 [59.7, 71.4] | 71.3 [65.9, 76.2] |
| Sex[c] | | N | 987 | 462 | 736 | 61,104 | 729 |
| | Female | n (%) | 520 (52.7%) | 226 (48.9%) | 329 (44.7%) | 32,326 (52.9%) | 488 (66.9%) |
| | Male | n (%) | 467 (47.3%) | 236 (51.1%) | 407 (55.3%) | 28,778 (47.1%) | 241 (33.1%) |
| Ethnicity | | N | 978 | – | 736 | 60,932 | 736 |
| | Asian | n (%) | 30 (3.1%) | – | 253 (34.4%) | 854 (1.4%) | < 10[d] |
| | Black | n (%) | 106 (10.8%) | – | 171 (23.2%) | 421 (0.7%) | 106 (14.4%) |
| | Mixed | n (%) | 21 (2.1%) | – | – | 321 (0.5%) | < 10[d] |
| | White | n (%) | 819 (83.7%) | – | 308 (41.8%) | 58,988 (96.8%) | 622 (84.5%) |
| | Other | n (%) | < 10[d] | – | < 10[d] | 348 (0.6%) | – |
| Total baseline WMH volume | | N | 1,007 | 462 | 765 | 61,104 | 844 |
| | | mL | 3.2 [1.4, 8.3] | 3.7 [1.9, 7.8] | 2.9 [1.6, 8.0] | 1.8 [1.1, 3.8] | 2.5 [1.3, 5.8] |
| APOE4 | | N | 796 | 462 | 736 | 57,298 | 732 |
| | None | n (%) | 495 (62.2%) | 323 (69.9%) | 617 (83.8%) | 41,522 (72.5%) | 437 (59.7%) |
| | 1 allele | n (%) | 250 (31.4%) | 127 (27.5%) | 106 (14.4%) | 14,540 (25.4%) | 254 (34.7%) |
| | 2 alleles | n (%) | 51 (6.4%) | 12 (2.6%) | 13 (1.8%) | 1,236 (2.2%) | 41 (5.6%) |
| Smoking | | N | – | 455 | 635 | 60,439 | 566 |
| | Current | n (%) | – | 16 (3.5%) | 22 (3.5%) | 2,023 (3.3%) | 36 (6.4%) |
| | Ever | n (%) | – | 281 (61.8%) | 219 (34.5%) | 20,380 (33.7%) | 226 (39.9%) |
| | Never | n (%) | – | 158 (34.7%) | 394 (62.0%) | 38,036 (62.9%) | 304 (53.7%) |
| Hypertension | | N | 987 | 22 | 736 | 61,104 | 451 |
| | No | n (%) | 555 (56.2%) | < 10[d] | 292 (39.7%) | 42,901 (70.2%) | 239 (53.0%) |
| | Yes | n (%) | 432 (43.8%) | 21 (95.5%) | 444 (60.3%) | 18,203 (29.8%) | 212 (47.0%) |
| Anti-hypertensive medication | | N | – | 462 | 736 | 60,360 | 736 |
| | No | n (%) | – | 275 (59.5%) | 310 (42.1%) | 45,331 (75.1%) | 483 (65.6%) |
| | Yes | n (%) | – | 187 (40.5%) | 426 (57.9%) | 15,029 (24.9%) | 253 (34.4%) |
| Systolic blood pressure | | N | 973 | 461 | 735 | 61,074 | 728 |
| | | mmHg | 133.0 [121.0, 145.0] | 137.5 [126.5, 148.0] | 140.0 [129.0, 152.5] | 135.0 [123.0, 148.0] | 130.0 [118.0, 140.0] |
| Diastolic blood pressure | | N | 973 | 461 | 735 | 61,074 | 703 |
| | | mmHg | 74.0 [68.0, 80.0] | 73.0 [66.5, 80.5] | 79.0 [72.0, 86.0] | 81.0 [74.0, 88.0] | 76.0 [70.0, 81.5] |
| Type 2 diabetes | | N | – | 462 | 736 | 61,104 | 452 |
| | No | n (%) | – | 411 (89.0%) | 641 (87.1%) | 58,558 (95.8%) | 403 (89.2%) |
| | Yes | n (%) | – | 51 (11.0%) | 95 (12.9%) | 2,546 (4.2%) | 49 (10.8%) |
| Diabetic medication | | N | – | 462 | 736 | 60,360 | 736 |
| | No | n (%) | – | 428 (92.6%) | 604 (82.1%) | 59,911 (99.3%) | 674 (91.6%) |
| | Yes | n (%) | – | 34 (7.4%) | 132 (17.9%) | 449 (0.7%) | 62 (8.4%) |
| HbA1c | | N | – | 462 | 719 | 56,727 | – |
| | | mmol/mol | – | 38.0 [36.0, 41.0] | 38.8 [34.7, 43.8] | 34.5 [32.2, 37.0] | – |
| Glucose | | N | 659 | 458 | 723 | 35,383 | – |
| | | mmol/L | 5.3 [5.0, 6.0] | 4.9 [4.4, 5.4] | 4.4 [4.0, 4.9] | 3.5 [3.1, 4.0] | – |
| Lipid-lowering medication | | N | – | 462 | 736 | 60,360 | 736 |
| | No | n (%) | – | 294 (63.6%) | 359 (48.8%) | 45,108 (74.7%) | 394 (53.5%) |
| | Yes | n (%) | – | 168 (36.4%) | 377 (51.2%) | 15,252 (25.3%) | 342 (46.5%) |
| LDL | | N | – | 462 | 721 | 35,445 | – |
| | | mmol/L | – | 3.0 [2.3, 3.8] | 2.4 [1.8, 3.1] | 1.7 [1.5, 2.0] | – |
| HDL | | N | – | 462 | 727 | 35,445 | – |
| | | mmol/L | – | 1.6 [1.3, 2.0] | 1.5 [1.3, 1.9] | 1.3 [1.1, 1.5] | – |
| Triglycerides | | N | 659 | 415 | 727 | 35,445 | – |
| | | mmol/L | 1.3 [1.0, 1.8] | 1.2 [0.9, 1.6] | 1.2 [0.9, 1.6] | 1.1 [0.8, 1.6] | – |
| WBCC | | N | – | 462 | 726 | 58,269 | – |
| | | $10^9$/L | – | 6.5 [5.5, 7.8] | 6.3 [5.3, 7.7] | 6.4 [5.4, 7.5] | – |
| Creatinine | | N | 659 | 458 | 727 | 34,565 | – |
| | | $\mu$mol/L | 80.4 [69.0, 93.7] | 73.0 [64.0, 84.0] | 78.0 [67.0, 91.0] | 65.7 [57.7, 74.6] | – |

a. Continuous variables reported as median [IQR].
b. For each variable, the preceding N row is the number of non-missing observations available in that cohort.
c. Categorical variables are reported as counts (%).
d. Categories with small counts were suppressed as $< 10$ to reduce disclosure risk.

#### e2.3 Comparison of internal dataset and OASIS-3

*eTable 7 Median and IQR of total WMH volume (mL) of each cluster in the internal dataset and OASIS-3.*

| DATASET | CLUSTER 1 | CLUSTER 2 | CLUSTER 3 | CLUSTER 4 | CLUSTER 5 |
|---|---|---|---|---|---|
| Internal | 0.9 [0.7, 1.3]mL | 1.5 [1.0, 2.1]mL | 1.9 [1.2, 3.0]mL | 4.4 [2.5, 7.8]mL | 6.4 [3.5, 11.9]mL |
| OASIS-3 | 0.8 [0.5, 1.2]mL | 1.5 [1.1, 2.4]mL | 1.7 [1.1, 3.0]mL | 5.2 [2.5, 10.5]mL | 6.0 [3.2, 11.9]mL |

#### e2.4 Identification of location patterns

The stability profiles of all clustering methods are shown in eFigure 6 below for k between 2 and 7. Using a stability threshold of 0.90, subKMeans achieved the highest stability of 0.946 when $k = 5$ ($0.944 \pm 0.001$ in 5-fold cross-validation), supporting the selection of $k = 5$ for subKMeans as the optimal number of clusters for the dataset. Representative participants from each location pattern are also shown.

*eFigure 6 Stability profiles of different clustering methods, averaged over 5-fold cross-validations.*

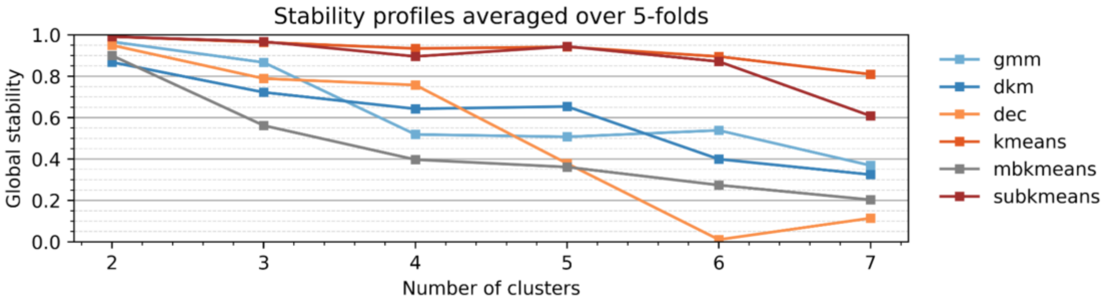


*eFigure 7 Proportions of (a) sex and (b) age groups (below 65, and 65 and above) in each cluster in the internal dataset.*

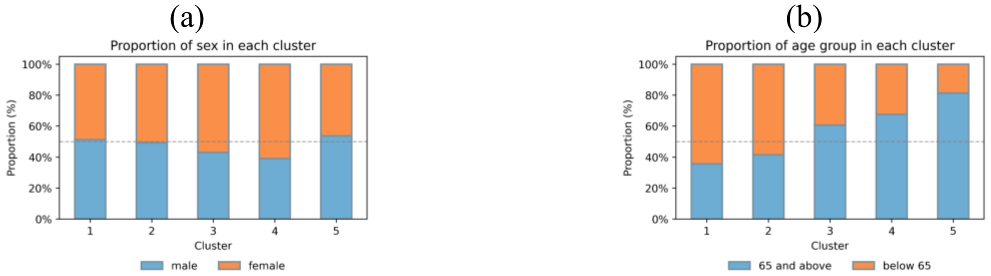


#### e2.5 Within-participant transitions between WMH location patterns

*eTable 8 Among individuals with 18-30 months follow-up interval, within-participant transitions in WMH location patterns between baseline and follow-up.*

| MOVEMENT TYPE | NUMBER OF PARTICIPANTS | PERCENT (%) |
|---|---|---|
| Stayed in same cluster | 2,697 | 71.5 |
| Moved within low-burden clusters 1-3 | 706 | 18.7 |
| Moved within high-burden clusters 4-5 | 59 | 1.6 |
| Moved 1/2/3 → 4/5 | 235 | 6.2 |
| Moved 4/5 → 1/2/3 | 75 | 2.0 |
| Total | 3,772 | 100.0 |

e2.6 Cross-sectional association between location patterns and risk factors

#### Internal dataset

*eTable 9 Cross-sectional associations between risk factors and proximity to centroids of 5 clusters the internal dataset, estimated using multivariable linear regression and adjusted by age, sex and total WMH volume. β is the standardised effect size, L and U are the lower and upper limits of the 95% CI. P-values were BH-corrected. ** indicates p < 0.001. Abbreviations: WMHV=total WMH volume, SBP/DBP=systolic/diastolic blood pressure, DM2=type 2 diabetes, MedChol=lipid-lowering medication, MedBP=anti-hypertensive medication, MedDM=diabetic medication, HDL=high-density lipoprotein cholesterol, LDL=low-density lipoprotein cholesterol, WBCC=white blood cell count. Categorical variables and reference categories: sex (male), smoking (never), ethnicity (white), number of APOE4 alleles (none), and diagnoses of medication conditions or use of medications (no).*

| RISK FACTOR | CATEGORY | CLUSTER 1 | | | | CLUSTER 2 | | | | CLUSTER 3 | | | | CLUSTER 4 | | | | CLUSTER 5 | | | |
|---|---|---|---|---|---|---|---|---|---|---|---|---|---|---|---|---|---|---|---|---|---|
| | | β | L | U | P-VALUE | β | L | U | P-VALUE | β | L | U | P-VALUE | β | L | U | P-VALUE | β | L | U | P-VALUE |
| WMHV | | -2.404 | -2.453 | -2.356 | ** | -1.816 | -1.875 | -1.758 | ** | -1.461 | -1.525 | -1.397 | ** | 2.792 | 2.731 | 2.854 | ** | 4.243 | 4.184 | 4.301 | ** |
| Age | | -0.446 | -0.462 | -0.431 | ** | -0.320 | -0.338 | -0.302 | ** | 0.203 | 0.183 | 0.223 | ** | 0.393 | 0.374 | 0.413 | ** | 0.545 | 0.527 | 0.564 | ** |
| Sex | Female | -0.012 | -0.016 | -0.007 | ** | -0.017 | -0.022 | -0.011 | ** | 0.040 | 0.035 | 0.046 | ** | 0.054 | 0.049 | 0.060 | ** | -0.015 | -0.021 | -0.010 | ** |
| Ethnicity | Other | 0.001 | -0.028 | 0.029 | 0.969 | -0.037 | -0.071 | -0.002 | 0.058 | -0.004 | -0.042 | 0.034 | 0.876 | -0.015 | -0.051 | 0.021 | 0.501 | -0.015 | -0.049 | 0.019 | 0.485 |
| Ethnicity | Mixed | -0.021 | -0.050 | 0.008 | 0.221 | -0.026 | -0.060 | 0.009 | 0.221 | -0.042 | -0.080 | -0.003 | 0.053 | -0.024 | -0.060 | 0.013 | 0.283 | -0.002 | -0.036 | 0.033 | 0.940 |
| Ethnicity | Asian | -0.028 | -0.044 | -0.011 | 0.002 | -0.054 | -0.074 | -0.034 | ** | -0.061 | -0.083 | -0.039 | ** | -0.039 | -0.060 | -0.019 | ** | -0.021 | -0.041 | -0.001 | 0.060 |
| Ethnicity | Black | -0.112 | -0.134 | -0.090 | ** | -0.073 | -0.099 | -0.047 | ** | -0.107 | -0.136 | -0.078 | ** | -0.013 | -0.041 | 0.014 | 0.442 | 0.048 | 0.022 | 0.074 | 0.001 |
| APOE4 | One | -0.003 | -0.008 | 0.002 | 0.295 | 0.003 | -0.003 | 0.009 | 0.422 | 0.001 | -0.006 | 0.008 | 0.814 | -0.001 | -0.007 | 0.006 | 0.902 | 0.001 | -0.005 | 0.007 | 0.711 |
| APOE4 | Two | -0.040 | -0.055 | -0.025 | ** | 0.013 | -0.005 | 0.031 | 0.221 | -0.048 | -0.067 | -0.028 | ** | -0.040 | -0.059 | -0.021 | ** | -0.009 | -0.026 | 0.009 | 0.440 |
| Smoking | Current | -0.021 | -0.033 | -0.009 | 0.002 | -0.011 | -0.026 | 0.003 | 0.195 | -0.004 | -0.020 | 0.012 | 0.696 | 0.004 | -0.011 | 0.019 | 0.678 | 0.021 | 0.006 | 0.035 | 0.009 |
| Smoking | Ever | -0.009 | -0.014 | -0.004 | ** | -0.009 | -0.015 | -0.004 | 0.002 | 0.004 | -0.002 | 0.010 | 0.254 | 0.002 | -0.004 | 0.008 | 0.589 | 0.004 | -0.002 | 0.009 | 0.221 |
| Hypertension | Yes | -0.033 | -0.037 | -0.028 | ** | -0.040 | -0.045 | -0.034 | ** | 0.013 | 0.007 | 0.020 | ** | 0.055 | 0.049 | 0.061 | ** | 0.031 | 0.025 | 0.037 | ** |
| MedBP | Yes | -0.031 | -0.036 | -0.026 | ** | -0.041 | -0.047 | -0.035 | ** | 0.013 | 0.007 | 0.020 | ** | 0.062 | 0.055 | 0.068 | ** | 0.033 | 0.027 | 0.039 | ** |
| DBP | | -0.091 | -0.111 | -0.071 | ** | -0.151 | -0.175 | -0.126 | ** | 0.121 | 0.094 | 0.148 | ** | 0.245 | 0.219 | 0.271 | ** | 0.164 | 0.139 | 0.188 | ** |
| SBP | | -0.101 | -0.123 | -0.079 | ** | -0.117 | -0.143 | -0.091 | ** | 0.091 | 0.062 | 0.120 | ** | 0.241 | 0.213 | 0.268 | ** | 0.166 | 0.140 | 0.193 | ** |
| DM2 | Yes | -0.035 | -0.045 | -0.024 | ** | -0.039 | -0.052 | -0.027 | ** | 0.020 | 0.006 | 0.034 | 0.008 | 0.015 | 0.002 | 0.029 | 0.042 | 0.003 | -0.010 | 0.015 | 0.719 |
| MedDM | Yes | -0.065 | -0.088 | -0.043 | ** | -0.041 | -0.068 | -0.014 | 0.005 | 0.006 | -0.024 | 0.035 | 0.764 | -0.011 | -0.039 | 0.018 | 0.538 | 0.015 | -0.012 | 0.042 | 0.367 |
| HbA1c | | -0.139 | -0.233 | -0.045 | 0.007 | -0.030 | -0.144 | 0.084 | 0.678 | -0.038 | -0.163 | 0.086 | 0.628 | 0.007 | -0.111 | 0.126 | 0.926 | 0.047 | -0.065 | 0.159 | 0.496 |
| Glucose | | -0.259 | -0.322 | -0.196 | ** | -0.065 | -0.140 | 0.011 | 0.144 | -0.063 | -0.146 | 0.020 | 0.205 | -0.089 | -0.168 | -0.010 | 0.046 | 0.076 | 0.001 | 0.151 | 0.077 |
| MedChol | Yes | -0.019 | -0.024 | -0.014 | ** | -0.032 | -0.038 | -0.025 | ** | 0.011 | 0.004 | 0.018 | 0.002 | 0.031 | 0.025 | 0.038 | ** | 0.009 | 0.003 | 0.015 | 0.009 |
| LDL | | -0.079 | -0.177 | 0.019 | 0.178 | 0.062 | -0.057 | 0.181 | 0.395 | -0.041 | -0.172 | 0.089 | 0.618 | -0.009 | -0.133 | 0.115 | 0.918 | 0.056 | -0.062 | 0.173 | 0.442 |
| HDL | | -0.079 | -0.190 | 0.031 | 0.221 | 0.096 | -0.037 | 0.229 | 0.221 | -0.061 | -0.207 | 0.084 | 0.496 | -0.036 | -0.175 | 0.102 | 0.678 | 0.061 | -0.070 | 0.192 | 0.449 |
| Triglycerides | | -0.114 | -0.161 | -0.067 | ** | -0.276 | -0.333 | -0.219 | ** | 0.081 | 0.018 | 0.143 | 0.020 | 0.157 | 0.098 | 0.217 | ** | -0.082 | -0.139 | -0.025 | 0.008 |
| Creatinine | | -0.246 | -0.321 | -0.171 | ** | -0.314 | -0.404 | -0.223 | ** | -0.238 | -0.338 | -0.138 | ** | -0.186 | -0.281 | -0.090 | ** | -0.235 | -0.325 | -0.144 | ** |
| WBCC | | -0.108 | -0.203 | -0.013 | 0.044 | 0.002 | -0.113 | 0.118 | 0.969 | -0.079 | -0.206 | 0.047 | 0.296 | 0.016 | -0.105 | 0.136 | 0.848 | 0.062 | -0.053 | 0.176 | 0.382 |

#### OASIS-3

*eTable 10 Cross-sectional associations between risk factors and proximity to centroids of 5 clusters the OASIS-3, estimated using multivariable linear regression and adjusted by age, sex and total WMH volume. β is the standardised effect size, L and U are the lower and upper limits of the 95% CI. P-values were BH-corrected. Abbreviations: WMHV=total WMH volume, SBP/DBP=systolic/diastolic blood pressure, DM2=type 2 diabetes, MedChol=lipid-lowering medication, MedBP=anti-hypertensive medication, MedDM=diabetic medication, HDL=high-density lipoprotein cholesterol, LDL=low-density lipoprotein cholesterol. Categorical variables and reference categories: sex (male), smoking (never), ethnicity (white), number of APOE4 alleles (none), and diagnoses of medication conditions or use of medications (no).*

| | | CLUSTER 1 | | | | CLUSTER 2 | | | | CLUSTER 3 | | | | CLUSTER 4 | | | | CLUSTER 5 | | | |
|---|---|---|---|---|---|---|---|---|---|---|---|---|---|---|---|---|---|---|---|---|---|
| **RISK FACTOR** | **CATEGORY** | **β** | **L** | **U** | **P-VALUE** | **β** | **L** | **U** | **P-VALUE** | **β** | **L** | **U** | **P-VALUE** | **β** | **L** | **U** | **P-VALUE** | **β** | **L** | **U** | **P-VALUE** |
| **WMHV** | | -0.681 | -0.835 | -0.528 | 0.000 | -0.992 | -1.193 | -0.791 | 0.000 | -0.873 | -1.087 | -0.658 | 0.000 | 0.538 | 0.362 | 0.714 | 0.000 | 0.831 | 0.587 | 1.074 | 0.000 |
| **Age** | | -0.285 | -0.372 | -0.197 | 0.000 | -0.161 | -0.276 | -0.046 | 0.039 | 0.000 | -0.123 | 0.123 | 1.000 | 0.226 | 0.125 | 0.326 | 0.000 | 0.652 | 0.513 | 0.791 | 0.000 |
| **Sex** | **Female** | -0.016 | -0.045 | 0.013 | 0.560 | 0.037 | -0.001 | 0.075 | 0.214 | -0.042 | -0.083 | -0.002 | 0.183 | -0.025 | -0.059 | 0.008 | 0.377 | 0.078 | 0.032 | 0.124 | 0.008 |
| **Ethnicity** | **Other** | | | | | | | | | | Insufficient participants | | | | | | | | | | |
| **Ethnicity** | **Mixed** | -0.125 | -0.336 | 0.085 | 0.521 | 0.003 | -0.272 | 0.278 | 1.000 | -0.148 | -0.444 | 0.147 | 0.588 | -0.163 | -0.407 | 0.081 | 0.451 | -0.292 | -0.628 | 0.044 | 0.285 |
| **Ethnicity** | **Asian** | -0.109 | -0.273 | 0.054 | 0.451 | -0.200 | -0.413 | 0.013 | 0.238 | -0.014 | -0.243 | 0.216 | 1.000 | -0.022 | -0.211 | 0.168 | 0.950 | -0.115 | -0.376 | 0.146 | 0.622 |
| **Ethnicity** | **Black** | -0.077 | -0.116 | -0.038 | 0.001 | -0.113 | -0.164 | -0.063 | 0.000 | -0.086 | -0.140 | -0.032 | 0.014 | -0.028 | -0.073 | 0.016 | 0.469 | -0.056 | -0.117 | 0.006 | 0.258 |
| **APOE4** | **One** | -0.028 | -0.057 | 0.001 | 0.214 | -0.002 | -0.041 | 0.036 | 1.000 | -0.015 | -0.055 | 0.026 | 0.727 | -0.003 | -0.037 | 0.030 | 0.969 | 0.023 | -0.023 | 0.070 | 0.588 |
| **APOE4** | **Two** | -0.086 | -0.147 | -0.026 | 0.036 | -0.022 | -0.101 | 0.058 | 0.813 | -0.104 | -0.188 | -0.019 | 0.085 | -0.017 | -0.087 | 0.052 | 0.839 | 0.101 | 0.005 | 0.197 | 0.183 |
| **Smoking** | **Current** | -0.012 | -0.078 | 0.055 | 0.866 | -0.042 | -0.129 | 0.044 | 0.588 | 0.068 | -0.022 | 0.159 | 0.377 | 0.077 | 0.000 | 0.154 | 0.199 | 0.038 | -0.065 | 0.142 | 0.727 |
| **Smoking** | **Ever** | 0.016 | -0.018 | 0.049 | 0.603 | -0.008 | -0.051 | 0.036 | 0.866 | 0.060 | 0.014 | 0.106 | 0.057 | 0.030 | -0.008 | 0.069 | 0.375 | 0.044 | -0.008 | 0.096 | 0.303 |
| **Hypertension** | **Yes** | -0.019 | -0.053 | 0.016 | 0.562 | -0.051 | -0.097 | -0.005 | 0.148 | -0.007 | -0.057 | 0.042 | 0.894 | 0.024 | -0.018 | 0.066 | 0.536 | 0.012 | -0.047 | 0.071 | 0.866 |
| **MedBP** | **Yes** | -0.009 | -0.038 | 0.020 | 0.774 | -0.012 | -0.050 | 0.026 | 0.774 | -0.007 | -0.048 | 0.033 | 0.866 | 0.000 | -0.033 | 0.034 | 1.000 | 0.025 | -0.021 | 0.071 | 0.560 |
| **DBP** | | -0.048 | -0.130 | 0.034 | 0.525 | 0.030 | -0.076 | 0.137 | 0.809 | -0.023 | -0.136 | 0.090 | 0.866 | 0.045 | -0.048 | 0.139 | 0.590 | 0.091 | -0.039 | 0.221 | 0.445 |
| **SBP** | | -0.080 | -0.185 | 0.025 | 0.377 | 0.005 | -0.133 | 0.142 | 1.000 | -0.035 | -0.181 | 0.112 | 0.842 | 0.027 | -0.094 | 0.147 | 0.854 | 0.172 | 0.006 | 0.338 | 0.183 |
| **DM2** | **Yes** | 0.010 | -0.044 | 0.063 | 0.866 | -0.019 | -0.091 | 0.052 | 0.813 | 0.036 | -0.040 | 0.112 | 0.597 | 0.044 | -0.020 | 0.109 | 0.451 | 0.030 | -0.062 | 0.122 | 0.774 |
| **MedDM** | **Yes** | 0.012 | -0.038 | 0.061 | 0.842 | -0.023 | -0.088 | 0.041 | 0.727 | 0.047 | -0.022 | 0.115 | 0.451 | 0.051 | -0.005 | 0.107 | 0.258 | 0.035 | -0.043 | 0.112 | 0.622 |
| **MedChol** | **Yes** | -0.014 | -0.041 | 0.014 | 0.588 | -0.011 | -0.048 | 0.025 | 0.774 | 0.025 | -0.014 | 0.064 | 0.467 | 0.042 | 0.010 | 0.073 | 0.057 | 0.028 | -0.016 | 0.072 | 0.469 |

### e2.7 Baseline characteristics of the longitudinal dataset

#### *eTable 11 Baseline characteristics comparing non-progression and progression groups in the Internal and OASIS-3 datasets.*

| | CATEGORY | UNIT | INTERNAL | | | OASIS-3 | | |
|---|---|---|---|---|---|---|---|---|
| | | | Non-progressor (n=3,779) | Progressor (n=1,495) | P-value[c] | Non-progressor (n=94) | Progressor (n=88) | P-value[c] |
| Age[a] | | years | 62.2 [55.8, 69.2] | 68.8 [62.1, 73.3] | < 0.001 | 65.5 [59.3, 70.6] | 71.7 [68.1, 76.5] | < 0.001 |
| Sex[b] | | | | | 0.739 | | | 0.629 |
| | Female | n | 1,997 (52.8%) | 779 (52.1%) | | 60 (63.8%) | 62 (70.5%) | |
| | Male | n | 1,782 (47.2%) | 716 (47.9%) | | 34 (36.2%) | 26 (29.5%) | |
| Ethnicity | | | | | 0.602 | | | 0.798 |
| | Asian | n | 44 (1.3%) | 17 (1.2%) | | < 10[d] | – | |
| | Black | n | 36 (1.0%) | 29 (2.0%) | | 10 (10.6%) | 11 (12.5%) | |
| | Mixed | n | 20 (0.6%) | 11 (0.7%) | | – | – | |
| | White | n | 3,324 (96.7%) | 1,412 (95.9%) | | 83 (88.3%) | 77 (87.5%) | |
| | Other | n | 14 (0.4%) | < 10[d] | | – | – | |
| Total baseline WMH volume | | mL | 1.4 [1.0, 2.4] | 3.8 [1.8, 7.2] | < 0.001 | 1.5 [0.9, 2.4] | 4.1 [2.5, 7.5] | 0.064 |
| dist1 | | – | 0.73 [0.63, 0.85] | 0.90 [0.76, 1.03] | < 0.001 | 0.90 [0.79, 1.01] | 1.01 [0.91, 1.13] | 0.049 |
| dist2 | | – | 0.71 [0.60, 0.84] | 0.79 [0.67, 0.93] | < 0.001 | 0.78 [0.67, 0.89] | 0.89 [0.77, 0.99] | 0.166 |
| dist3 | | – | 0.74 [0.63, 0.86] | 0.75 [0.62, 0.90] | 0.858 | 0.80 [0.69, 0.93] | 0.83 [0.72, 0.96] | 0.820 |
| dist4 | | – | 0.89 [0.75, 1.01] | 0.72 [0.60, 0.86] | < 0.001 | 0.91 [0.79, 1.01] | 0.81 [0.74, 0.90] | 0.080 |
| dist5 | | – | 0.94 [0.79, 1.07] | 0.72 [0.58, 0.88] | < 0.001 | 0.93 [0.76, 1.02] | 0.72 [0.59, 0.82] | < 0.001 |
| APOE4 | | | | | 0.005 | | | 0.347 |
| | None | n | 2,570 (72.4%) | 949 (69.2%) | | 65 (69.9%) | 58 (65.9%) | |
| | 1 allele | n | 911 (25.6%) | 369 (26.9%) | | 27 (29.0%) | 24 (27.3%) | |
| | 2 alleles | n | 71 (2.0%) | 53 (3.9%) | | < 10[d] | < 10[d] | |
| Smoking | | | | | 0.858 | | | 0.925 |
| | Current | n | 121 (3.5%) | 36 (3.2%) | | < 10[d] | < 10[d] | |
| | Ever | n | 1,171 (33.4%) | 419 (37.2%) | | 26 (31.7%) | 28 (41.8%) | |
| | Never | n | 2,212 (63.1%) | 671 (59.6%) | | 49 (59.8%) | 33 (49.3%) | |
| Hypertension | | | | | 0.004 | | | 0.518 |
| | No | n | 2,620 (75.7%) | 938 (63.4%) | | 14 (77.8%) | 16 (51.6%) | |
| | Yes | n | 841 (24.3%) | 541 (36.6%) | | < 10[d] | 15 (48.4%) | |
| Anti-hypertensive medication | | | | | 0.156 | | | 0.489 |
| | No | n | 2,833 (80.8%) | 828 (73.3%) | | 71 (75.5%) | 52 (59.1%) | |
| | Yes | n | 672 (19.2%) | 302 (26.7%) | | 23 (24.5%) | 36 (40.9%) | |
| Systolic blood pressure | | mmHg | 132.0 [121.0, 144.0] | 138.0 [126.0, 150.0] | < 0.001 | 124.0 [117.2, 135.8] | 130.0 [118.0, 139.2] | 0.518 |
| Diastolic blood pressure | | mmHg | 79.0 [72.0, 86.0] | 80.0 [73.0, 88.0] | < 0.001 | 76.0 [70.0, 80.0] | 76.0 [70.0, 81.8] | 0.347 |
| Type 2 diabetes | | | | | 0.189 | | | 0.418 |
| | No | n | 3,417 (96.9%) | 1,083 (95.2%) | | 14 (77.8%) | 29 (93.5%) | |
| | Yes | n | 111 (3.1%) | 55 (4.8%) | | < 10[d] | < 10[d] | |
| Diabetic medication | | | | | 0.189 | | | 0.418 |
| | No | n | 3,464 (98.8%) | 1,121 (99.2%) | | 85 (90.4%) | 81 (92.0%) | |
| | Yes | n | 41 (1.2%) | < 10[d] | | < 10[d] | < 10[d] | |
| HbA1c | | mmol/mol | 34.300 [32.000, 36.900] | 34.800 [32.600, 37.400] | < 0.001 | – | – | – |
| Glucose | | mmol/L | 3.644 [3.171, 4.300] | 3.819 [3.278, 4.799] | 0.215 | – | – | – |
| Lipid-lowering medication | | | | | 0.006 | | | 0.820 |
| | No | n | 2,884 (82.3%) | 809 (71.6%) | | 63 (67.0%) | 50 (56.8%) | |
| | Yes | n | 621 (17.7%) | 321 (28.4%) | | 31 (33.0%) | 38 (43.2%) | |
| HDL | | mmol/L | 1.350 [1.128, 1.600] | 1.295 [1.116, 1.552] | < 0.001 | – | – | – |
| LDL | | mmol/L | 1.797 [1.501, 2.160] | 1.739 [1.482, 2.050] | < 0.001 | – | – | – |
| Triglycerides | | mmol/L | 1.135 [0.845, 1.586] | 1.227 [0.854, 1.631] | 0.858 | – | – | – |
| White blood cell count | | $10^9$/L | 6.330 [5.400, 7.500] | 6.330 [5.400, 7.470] | < 0.001 | – | – | – |
| Creatinine | | $\mu$mol/L | 67.596 [58.732, 77.555] | 68.776 [59.481, 79.663] | 0.156 | – | – | – |

a. For continuous variables, Median [IQR] are provided and between-group differences were compared using Mann-Whitney U test after removing the effects of age and sex.
b. For categorical variables, counts of participants (%) are provided, and between-group differences were compared using Chi-squared test after removing the effects of age and sex.
c. P-values were BH-corrected in internal dataset and OASIS-3 respectively.
d. Categories with small counts suppressed as < 10.

#### e2.8 Longitudinal prediction

*eTable 12 Model performance for WMH progression prediction, reported as balanced accuracy (mean and 95% CI) across train, test, and OASIS-3 datasets. Abbreviations: clusters = spatial patterns of WMH, distance = Euclidean distance to five WMH cluster centroids, RF = age, sex, and systolic blood pressure, WMHV = total baseline WMH volume, RWMHV = 36 regional baseline WMH volume, PCV = principal components of RWMHV.*

| MODEL | TRAIN (95% CI) | | | TEST (95% CI) | | | OASIS-3 (95% CI) | | |
|---|---|---|---|---|---|---|---|---|---|
| | MEAN | LOWER | UPPER | MEAN | LOWER | UPPER | MEAN | LOWER | UPPER |
| RF | 0.632 | 0.617 | 0.646 | 0.657 | 0.626 | 0.686 | 0.687 | 0.622 | 0.751 |
| WMHV | 0.707 | 0.680 | 0.734 | 0.693 | 0.664 | 0.723 | 0.748 | 0.688 | 0.804 |
| PCV | 0.724 | 0.704 | 0.744 | 0.693 | 0.662 | 0.722 | 0.777 | 0.718 | 0.831 |
| clusters | 0.696 | 0.680 | 0.712 | 0.649 | 0.619 | 0.677 | 0.689 | 0.626 | 0.750 |
| distance | 0.729 | 0.712 | 0.747 | 0.712 | 0.681 | 0.742 | 0.711 | 0.653 | 0.769 |
| WMHV + RF | 0.729 | 0.706 | 0.751 | 0.706 | 0.676 | 0.735 | 0.778 | 0.717 | 0.840 |
| PCV + RF | 0.737 | 0.713 | 0.762 | 0.702 | 0.670 | 0.730 | 0.785 | 0.727 | 0.846 |
| clusters + RF | 0.716 | 0.690 | 0.742 | 0.700 | 0.669 | 0.730 | 0.697 | 0.635 | 0.759 |
| distance + RF | 0.735 | 0.699 | 0.772 | 0.709 | 0.678 | 0.738 | 0.706 | 0.647 | 0.769 |
| WMHV + PCV | 0.733 | 0.708 | 0.758 | 0.695 | 0.664 | 0.724 | 0.795 | 0.736 | 0.848 |
| WMHV + clusters | 0.720 | 0.703 | 0.738 | 0.692 | 0.662 | 0.722 | 0.752 | 0.689 | 0.813 |
| WMHV + distance | 0.736 | 0.718 | 0.754 | 0.712 | 0.680 | 0.742 | 0.727 | 0.664 | 0.787 |
| WMHV + PCV + RF | 0.739 | 0.721 | 0.757 | 0.710 | 0.679 | 0.740 | 0.791 | 0.735 | 0.849 |
| WMHV + clusters + RF | 0.736 | 0.711 | 0.760 | 0.701 | 0.669 | 0.730 | 0.752 | 0.694 | 0.815 |
| WMHV + distance + RF | 0.743 | 0.724 | 0.763 | 0.707 | 0.677 | 0.736 | 0.732 | 0.669 | 0.790 |
| RWMHV | 0.745 | 0.730 | 0.760 | 0.733 | 0.703 | 0.762 | 0.741 | 0.676 | 0.801 |
| RWMHV + RF | 0.749 | 0.742 | 0.756 | 0.730 | 0.701 | 0.757 | 0.741 | 0.678 | 0.801 |
| RWMHV + PCV | 0.745 | 0.734 | 0.756 | 0.728 | 0.699 | 0.756 | 0.757 | 0.695 | 0.814 |
| RWMHV + clusters | 0.753 | 0.734 | 0.772 | 0.733 | 0.702 | 0.760 | 0.741 | 0.677 | 0.802 |
| RWMHV + distance | 0.742 | 0.728 | 0.757 | 0.734 | 0.704 | 0.761 | 0.741 | 0.678 | 0.798 |
| RWMHV + PCV + RF | 0.748 | 0.739 | 0.757 | 0.737 | 0.706 | 0.764 | 0.764 | 0.705 | 0.821 |
| RWMHV + clusters + RF | 0.745 | 0.717 | 0.773 | 0.725 | 0.695 | 0.752 | 0.725 | 0.659 | 0.791 |
| RWMHV + distance + RF | 0.748 | 0.736 | 0.759 | 0.736 | 0.707 | 0.762 | 0.741 | 0.674 | 0.800 |

#### Internal dataset

*eTable 13 BH-corrected p-values from pairwise McNemar's tests between models for WMH progression in internal test set. bold indicates p-values < 0.05, ** indicates p-values < 0.001. Abbreviations: clusters=location patterns of WMH, distance=Euclidean distance to five cluster centroids, RF=age, sex and systolic blood pressure, WMHV=total baseline WMH volume, RWMHV=36 regional baseline WMH volume, PCV=principal components of RWMHV.*

| | RF | WMHV | PCV | CLUSTERS | DISTANCE | WMHV + RF | PCV + RF | CLUSTERS + RF | DISTANCE + RF | WMHV + PCV | WMHV + CLUSTERS | WMHV + DISTANCE | WMHV + PCV + RF | WMHV + CLUSTERS + RF | WMHV + DISTANCE + RF | RWMHV | RWMHV + RF | RWMHV + PCV | RWMHV + CLUSTERS | RWMHV + DISTANCE | RWMHV + PCV + RF | RWMHV + CLUSTERS + RF | RWMHV + DISTANCE + RF |
|---|---|---|---|---|---|---|---|---|---|---|---|---|---|---|---|---|---|---|---|---|---|---|---|
| **RF** | | **0.015** | **0.030** | **0.027** | **0.025** | ** | **0.005** | 0.148 | **0.022** | **0.026** | 0.076 | **0.004** | **0.002** | **0.003** | **0.006** | ** | ** | ** | ** | ** | ** | ** | ** |
| **WMHV** | **0.016** | | 0.664 | ** | 0.816 | 0.136 | 0.973 | 0.168 | 0.763 | 0.771 | 0.334 | 0.763 | 0.763 | 1.000 | 1.000 | **0.023** | **0.046** | 0.089 | 0.060 | 0.100 | **0.046** | 0.124 | 0.089 |
| **PCV** | **0.030** | 0.664 | | ** | 1.000 | **0.042** | 0.454 | 0.292 | 1.000 | 0.938 | 0.694 | 0.369 | 0.252 | 0.664 | 0.669 | **0.002** | **0.006** | **0.011** | **0.008** | **0.017** | **0.004** | **0.022** | **0.017** |
| **CLUSTERS** | **0.027** | ** | ** | | ** | ** | ** | ** | ** | ** | ** | ** | ** | ** | ** | ** | ** | ** | ** | ** | ** | ** | ** |
| **DISTANCE** | **0.025** | 0.816 | 1.000 | ** | | 0.160 | 0.712 | 0.242 | 0.973 | 1.000 | 0.709 | 0.296 | 0.536 | 0.763 | 0.709 | **0.006** | **0.009** | **0.026** | **0.011** | **0.014** | **0.009** | **0.027** | **0.017** |
| **WMHV + RF** | ** | 0.136 | **0.042** | ** | 0.160 | | 0.116 | **0.006** | 0.110 | 0.076 | **0.027** | 0.559 | 0.297 | 0.093 | 0.176 | 0.263 | 0.370 | 0.669 | 0.490 | 0.646 | 0.388 | 0.704 | 0.587 |
| **PCV + RF** | **0.005** | 0.973 | 0.454 | ** | 0.712 | 0.116 | | 0.057 | 0.604 | 0.631 | 0.296 | 0.816 | 0.669 | 1.000 | 1.000 | **0.021** | **0.026** | 0.071 | **0.045** | 0.082 | **0.018** | 0.081 | 0.068 |
| **CLUSTERS + RF** | 0.148 | 0.168 | 0.292 | ** | 0.242 | **0.006** | 0.057 | | 0.223 | 0.247 | 0.560 | **0.040** | **0.031** | **0.042** | 0.055 | ** | ** | ** | ** | ** | ** | ** | ** |
| **DISTANCE + RF** | **0.022** | 0.763 | 1.000 | ** | 0.973 | 0.110 | 0.604 | 0.223 | | 0.958 | 0.808 | 0.308 | 0.405 | 0.656 | 0.567 | **0.004** | **0.004** | **0.019** | **0.007** | **0.014** | **0.004** | **0.014** | **0.008** |
| **WMHV + PCV** | **0.026** | 0.771 | 0.938 | ** | 1.000 | 0.076 | 0.631 | 0.247 | 0.958 | | 0.587 | 0.481 | 0.340 | 0.763 | 0.768 | **0.005** | **0.011** | **0.019** | **0.016** | **0.030** | **0.007** | **0.037** | **0.030** |
| **WMHV + CLUSTERS** | 0.076 | 0.334 | 0.694 | ** | 0.709 | **0.027** | 0.296 | 0.560 | 0.808 | 0.587 | | 0.121 | 0.176 | 0.323 | 0.353 | ** | **0.004** | **0.006** | **0.004** | **0.008** | **0.003** | **0.013** | **0.008** |
| **WMHV + DISTANCE** | **0.004** | 0.763 | 0.369 | ** | 0.296 | 0.559 | 0.816 | **0.040** | 0.308 | 0.481 | 0.121 | | 1.000 | 0.775 | 0.709 | **0.038** | 0.075 | 0.155 | 0.082 | 0.118 | 0.075 | 0.191 | 0.124 |
| **WMHV + PCV + RF** | **0.002** | 0.763 | 0.252 | ** | 0.536 | 0.297 | 0.669 | **0.031** | 0.405 | 0.340 | 0.176 | 1.000 | | 0.771 | 0.784 | **0.038** | **0.046** | 0.135 | 0.081 | 0.143 | **0.030** | 0.148 | 0.117 |
| **WMHV + CLUSTERS + RF** | **0.004** | 1.000 | 0.664 | ** | 0.763 | 0.093 | 1.000 | **0.042** | 0.656 | 0.763 | 0.323 | 0.775 | 0.771 | | 0.973 | **0.021** | **0.027** | 0.081 | **0.044** | 0.082 | **0.024** | 0.082 | 0.064 |
| **WMHV + DISTANCE + RF** | **0.006** | 1.000 | 0.669 | ** | 0.709 | 0.176 | 1.000 | 0.055 | 0.567 | 0.768 | 0.353 | 0.709 | 0.784 | 0.973 | | **0.017** | **0.022** | 0.067 | **0.037** | 0.055 | **0.021** | 0.061 | **0.040** |
| **RWMHV** | ** | **0.023** | **0.002** | ** | **0.006** | 0.263 | **0.021** | ** | **0.004** | **0.005** | ** | **0.038** | **0.038** | **0.021** | **0.017** | | 0.810 | 0.413 | 0.676 | 0.454 | 0.776 | 0.370 | 0.588 |
| **RWMHV + RF** | ** | **0.046** | **0.006** | ** | **0.009** | 0.370 | **0.026** | ** | **0.004** | **0.011** | **0.004** | 0.075 | **0.046** | **0.027** | **0.022** | 0.810 | | 0.692 | 0.970 | 0.763 | 1.000 | 0.516 | 0.808 |
| **RWMHV + PCV** | ** | 0.089 | **0.011** | ** | **0.026** | 0.669 | 0.071 | ** | **0.019** | **0.019** | **0.006** | 0.155 | 0.135 | 0.081 | 0.067 | 0.413 | 0.692 | | 0.809 | 1.000 | 0.737 | 1.000 | 0.973 |
| **RWMHV + CLUSTERS** | ** | 0.060 | **0.008** | ** | **0.011** | 0.490 | **0.045** | ** | **0.007** | **0.016** | **0.004** | 0.082 | 0.081 | **0.044** | **0.037** | 0.676 | 0.970 | 0.809 | | 0.881 | 1.000 | 0.733 | 0.970 |
| **RWMHV + DISTANCE** | ** | 0.100 | **0.017** | ** | **0.014** | 0.646 | 0.082 | ** | **0.014** | **0.030** | **0.008** | 0.118 | 0.143 | 0.082 | 0.055 | 0.454 | 0.763 | 1.000 | 0.881 | | 0.834 | 0.970 | 1.000 |
| **RWMHV + PCV + RF** | ** | **0.046** | **0.004** | ** | **0.009** | 0.388 | **0.018** | ** | **0.004** | **0.007** | **0.003** | 0.075 | **0.030** | **0.024** | **0.021** | 0.776 | 1.000 | 0.737 | 1.000 | 0.834 | | 0.689 | 0.893 |
| **RWMHV + CLUSTERS + RF** | ** | 0.124 | **0.022** | ** | **0.027** | 0.704 | 0.081 | ** | **0.014** | **0.037** | **0.013** | 0.191 | 0.148 | 0.082 | 0.061 | 0.370 | 0.516 | 1.000 | 0.733 | 0.970 | 0.689 | | 0.881 |
| **RWMHV + DISTANCE + RF** | ** | 0.089 | **0.017** | ** | **0.017** | 0.587 | 0.068 | ** | **0.008** | **0.030** | **0.008** | 0.124 | 0.117 | 0.064 | **0.040** | 0.588 | 0.808 | 0.973 | 0.970 | 1.000 | 0.893 | 0.881 | |

#### OASIS-3

*eTable 14 BH-corrected p-values from pairwise McNemar's tests between models for WMH progression in OASIS-3. bold indicates p-values < 0.05, ** indicates p-values < 0.001. Abbreviations: clusters=location patterns of WMH, distance=Euclidean distance to five cluster centroids, RF=age, sex and systolic blood pressure, WMHV=total baseline WMH volume, RWMHV=36 regional baseline WMH volume, PCV=principal components of RWMHV.*

| | RF | WMHV | PCV | CLUSTERS | DISTANCE | WMHV + RF | PCV + RF | CLUSTERS + RF | DISTANCE + RF | WMHV + PCV | WMHV + CLUSTERS | WMHV + DISTANCE | WMHV + PCV + RF | WMHV + CLUSTERS + RF | WMHV + DISTANCE + RF | RWMHV | RWMHV + RF | RWMHV + PCV | RWMHV + CLUSTERS | RWMHV + DISTANCE | RWMHV + PCV + RF | RWMHV + CLUSTERS + RF | RWMHV + DISTANCE + RF |
|---|---|---|---|---|---|---|---|---|---|---|---|---|---|---|---|---|---|---|---|---|---|---|---|
| **RF** | | 0.556 | 0.317 | 0.945 | 0.890 | 0.227 | 0.213 | 0.922 | 0.922 | 0.213 | 0.523 | 0.688 | 0.213 | 0.393 | 0.620 | 0.588 | 0.588 | 0.469 | 0.588 | 0.588 | 0.342 | 0.661 | 0.584 |
| **WMHV** | 0.556 | | 0.636 | 0.504 | 0.635 | 0.677 | 0.622 | 0.584 | 0.616 | 0.435 | 0.922 | 0.801 | 0.588 | 0.922 | 0.868 | 0.922 | 0.922 | 1.000 | 0.922 | 0.922 | 0.922 | 0.788 | 0.922 |
| **PCV** | 0.317 | 0.636 | | 0.227 | 0.355 | 0.922 | 1.000 | 0.307 | 0.325 | 0.818 | 0.724 | 0.471 | 0.922 | 0.742 | 0.504 | 0.620 | 0.620 | 0.792 | 0.620 | 0.620 | 0.899 | 0.453 | 0.610 |
| **CLUSTERS** | 0.945 | 0.504 | 0.227 | | 0.868 | 0.227 | 0.213 | 0.922 | 0.922 | 0.213 | 0.317 | 0.636 | 0.213 | 0.445 | 0.616 | 0.588 | 0.588 | 0.423 | 0.584 | 0.588 | 0.342 | 0.644 | 0.556 |
| **DISTANCE** | 0.890 | 0.635 | 0.355 | 0.868 | | 0.342 | 0.227 | 0.922 | 1.000 | 0.213 | 0.610 | 0.849 | 0.213 | 0.588 | 0.763 | 0.648 | 0.648 | 0.489 | 0.641 | 0.641 | 0.393 | 0.909 | 0.648 |
| **WMHV + RF** | 0.227 | 0.677 | 0.922 | 0.227 | 0.342 | | 1.000 | 0.213 | 0.302 | 0.890 | 0.742 | 0.453 | 0.922 | 0.588 | 0.435 | 0.635 | 0.620 | 0.849 | 0.636 | 0.635 | 0.890 | 0.445 | 0.610 |
| **PCV + RF** | 0.213 | 0.622 | 1.000 | 0.213 | 0.227 | 1.000 | | 0.213 | 0.213 | 0.922 | 0.636 | 0.339 | 1.000 | 0.588 | 0.339 | 0.588 | 0.584 | 0.724 | 0.588 | 0.588 | 0.742 | 0.317 | 0.504 |
| **CLUSTERS + RF** | 0.922 | 0.584 | 0.307 | 0.922 | 0.922 | 0.213 | 0.213 | | 1.000 | 0.213 | 0.453 | 0.784 | 0.213 | 0.339 | 0.648 | 0.627 | 0.620 | 0.489 | 0.621 | 0.620 | 0.375 | 0.769 | 0.588 |
| **DISTANCE + RF** | 0.922 | 0.616 | 0.325 | 0.922 | 1.000 | 0.302 | 0.213 | 1.000 | | 0.213 | 0.572 | 0.742 | 0.213 | 0.504 | 0.636 | 0.620 | 0.620 | 0.445 | 0.616 | 0.616 | 0.355 | 0.849 | 0.616 |
| **WMHV + PCV** | 0.213 | 0.435 | 0.818 | 0.213 | 0.213 | 0.890 | 0.922 | 0.213 | 0.213 | | 0.453 | 0.227 | 1.000 | 0.504 | 0.302 | 0.471 | 0.445 | 0.584 | 0.471 | 0.453 | 0.620 | 0.227 | 0.393 |
| **WMHV + CLUSTERS** | 0.523 | 0.922 | 0.724 | 0.317 | 0.610 | 0.742 | 0.636 | 0.453 | 0.572 | 0.453 | | 0.761 | 0.584 | 0.922 | 0.849 | 0.922 | 0.922 | 1.000 | 0.922 | 0.922 | 0.922 | 0.742 | 0.922 |
| **WMHV + DISTANCE** | 0.688 | 0.801 | 0.471 | 0.636 | 0.849 | 0.453 | 0.339 | 0.784 | 0.742 | 0.227 | 0.761 | | 0.227 | 0.742 | 1.000 | 0.899 | 0.896 | 0.620 | 0.890 | 0.868 | 0.556 | 0.922 | 0.896 |
| **WMHV + PCV + RF** | 0.213 | 0.588 | 0.922 | 0.213 | 0.213 | 0.922 | 1.000 | 0.213 | 0.213 | 1.000 | 0.584 | 0.227 | | 0.489 | 0.227 | 0.523 | 0.489 | 0.622 | 0.556 | 0.523 | 0.621 | 0.227 | 0.435 |
| **WMHV + CLUSTERS + RF** | 0.393 | 0.922 | 0.742 | 0.445 | 0.588 | 0.588 | 0.588 | 0.339 | 0.504 | 0.504 | 0.922 | 0.742 | 0.489 | | 0.784 | 0.922 | 0.922 | 1.000 | 0.922 | 0.922 | 0.922 | 0.724 | 0.922 |
| **WMHV + DISTANCE + RF** | 0.620 | 0.868 | 0.504 | 0.616 | 0.763 | 0.435 | 0.339 | 0.648 | 0.636 | 0.302 | 0.849 | 1.000 | 0.227 | 0.784 | | 0.922 | 0.922 | 0.742 | 0.922 | 0.922 | 0.616 | 1.000 | 0.922 |
| **RWMHV** | 0.588 | 0.922 | 0.620 | 0.588 | 0.648 | 0.635 | 0.588 | 0.627 | 0.620 | 0.471 | 0.922 | 0.899 | 0.523 | 0.922 | 0.922 | | 0.899 | 0.861 | 0.922 | 0.922 | 0.763 | 0.884 | 0.922 |
| **RWMHV + RF** | 0.588 | 0.922 | 0.620 | 0.588 | 0.648 | 0.620 | 0.584 | 0.620 | 0.620 | 0.445 | 0.922 | 0.896 | 0.489 | 0.922 | 0.922 | 0.899 | | 0.849 | 0.922 | 0.922 | 0.724 | 0.861 | 0.922 |
| **RWMHV + PCV** | 0.469 | 1.000 | 0.792 | 0.423 | 0.489 | 0.849 | 0.724 | 0.489 | 0.445 | 0.584 | 1.000 | 0.620 | 0.622 | 1.000 | 0.742 | 0.861 | 0.849 | | 0.861 | 0.861 | 1.000 | 0.604 | 0.868 |
| **RWMHV + CLUSTERS** | 0.588 | 0.922 | 0.620 | 0.584 | 0.641 | 0.636 | 0.588 | 0.621 | 0.616 | 0.471 | 0.922 | 0.890 | 0.556 | 0.922 | 0.922 | 0.922 | 0.922 | 0.861 | | 0.922 | 0.792 | 0.884 | 0.922 |
| **RWMHV + DISTANCE** | 0.588 | 0.922 | 0.620 | 0.588 | 0.641 | 0.635 | 0.588 | 0.620 | 0.616 | 0.453 | 0.922 | 0.868 | 0.523 | 0.922 | 0.922 | 0.922 | 0.922 | 0.861 | 0.922 | | 0.792 | 0.868 | 0.922 |
| **RWMHV + PCV + RF** | 0.342 | 0.922 | 0.899 | 0.342 | 0.393 | 0.890 | 0.742 | 0.375 | 0.355 | 0.620 | 0.922 | 0.556 | 0.621 | 0.922 | 0.616 | 0.763 | 0.724 | 1.000 | 0.792 | 0.792 | | 0.523 | 0.742 |
| **RWMHV + CLUSTERS + RF** | 0.661 | 0.788 | 0.453 | 0.644 | 0.909 | 0.445 | 0.317 | 0.769 | 0.849 | 0.227 | 0.742 | 0.922 | 0.227 | 0.724 | 1.000 | 0.884 | 0.861 | 0.604 | 0.884 | 0.868 | 0.523 | | 0.784 |
| **RWMHV + DISTANCE + RF** | 0.584 | 0.922 | 0.610 | 0.556 | 0.648 | 0.610 | 0.504 | 0.588 | 0.616 | 0.393 | 0.922 | 0.896 | 0.435 | 0.922 | 0.922 | 0.922 | 0.922 | 0.868 | 0.922 | 0.922 | 0.742 | 0.784 | |
